# Synaptic resilience is associated with maintained cognition during ageing

**DOI:** 10.1101/2022.05.04.22274679

**Authors:** Declan King, Kris Holt, Jamie Toombs, Xin He, Owen Dando, J. A. Okely, Jamie Rose, Ciaran Gunn, Adele Correia, Carmen Montero, Jane Tulloch, Douglas Lamont, Adele M Taylor, Sarah E Harris, Paul Redmond, Simon R Cox, Christopher M Henstridge, Ian J Deary, Colin Smith, Tara L Spires-Jones

**Author notes:** **Correspondence to** Prof Tara Spires-Jones, UK Dementia Research Institute and Centre for Discovery Brain Sciences at the University of Edinburgh, 1 Goerge Square, Edinburgh, UK EH8 9JZ. **DECLARATION OF INTERESTS** TSJ received collaborative grant funding from an anonymous industry partner for this work. TSJ is on the scientific advisory board of Cognition Therapeutics, receives honoraria for talks from academic and industry, and is a trustee of the Guarantors of Brain and the British Neuroscience Association.

## Abstract

**INTRODUCTION:** It remains unclear why age increases risk of Alzheimer’s disease and why some people experience age-related cognitive decline in the absence of dementia. Here we test the hypothesis that resilience to molecular changes in synapses contribute to healthy cognitive ageing.

**METHODS:** We examined post-mortem brain from people in mid-life (n=15), healthy ageing with either maintained cognition (n=8) or lifetime cognitive decline (n=7), and Alzheimer’s disease (n=13). Synapses were examined with high resolution imaging, proteomics, and RNA sequencing. Stem cell-derived neurons were challenged with Alzheimer’s brain homogenate.

**RESULTS:** Synaptic pathology increased, and expression of genes involved in synaptic signalling decreased between mid-life, healthy ageing and Alzheimer’s. In contrast, brain tissue and neurons from people with maintained cognition during ageing exhibited decreases in synaptic signalling genes compared to people with cognitive decline.

**DISCUSSION:** Efficient synaptic networks without pathological protein accumulation may contribute to maintained cognition during ageing.

## BACKGROUND

Cognitive decline is described as one of the most feared aspects of the ageing process [1-4]. As well as cognitive decline being common during ageing, age is the most important risk factor for Alzheimer’s disease (AD). Region-specific synapse loss has been observed in post-mortem studies of aged human brain and in animals similar changes are associated with cognitive decline [5]. In AD, synapse loss also correlates strongly with cognitive decline [6-8] and we have observed that synaptic accumulation of pathological forms of amyloid beta (Aβ) and tau are associated with synapse loss in AD brain [9-11]. In model systems, we and others have observed that altered synaptic signalling downstream of Aβ and tau cause cognitive decline in ageing animals [12]. Several well-characterized cohorts have been used to study brain changes associated with cognitive ageing including the Religious Orders Study, Rush Memory and Ageing Project and the Cognitive Function and Ageing Studies [13-16]. These and other studies highlight the importance of different responses to pathological protein accumulation and both the genetic and environmental risk factors associated with age-related cognitive decline. However, significant gaps in knowledge remain including understanding brain changes associated with cognitive change over a large portion of the lifetime (starting in childhood) and detailed analysis of synapse density and protein composition at high resolution previously prevented by technical limitations. Here we address some of these knowledge gaps using a combination of advanced techniques and brain donations from the participants in the Lothian Birth Cohort 1936 (LBC1936), a well-characterised cohort studying cognitive ageing [17-21]. LBC1936 participants took a version of the Moray House Test No. 12 (MHT) of general intelligence at age 11 and have participated since the age of 70 in a longitudinal study of cognitive ageing. Using brain samples and induced pluripotent stem cell (iPSC) derived neurons derived from this unique cohort alongside brain tissue donated from middle-aged people who died from non-neurological conditions, and people who died with AD [22, 23], we have conducted an in-depth study of synaptic pathology and molecular composition to study synaptic changes associated with resilience to cognitive decline in ageing.

## METHODS

Full methods details can be found in the Supplementary Methods File. Abbreviated methods follow.

### Subjects

Brain tissue and blood donations have been reviewed and approved for use by the Edinburgh Brain Bank ethics committee and the Academic and Clinical Central Office for Research and Development, a joint office of the University of Edinburgh and NHS Lothian (approval 15-HV-016). Three groups of participants were included: (1) mid-life controls (age range 19-58, n=15); (2) healthy agers from LBC1936 (age range 77-84, n=16) and (3) Alzheimer’s disease (age range 61-95, n=13). Table 1 shows summary demographics. Detailed information for each participant is included in DOI Supplementary Methods Table 1.

**Table 1:** Summary demographic data of human brain tissue donors

| **Age (Years)** | **Midlife (ML)**<br>**(N=15)** | **Healthy Agers (HA)\***<br>**(N=16)** | **Alzheimer's Disease (AD)**<br>**(N=13)** | **Overall**<br>**(N=44)** |
| --- | --- | --- | --- | --- |
| Mean (SD) | 42.7 (9.41) | 80.2 (1.97) | 80.8 (11.0) | 67.6 (19.9) |
| Median (Min, Max) | 46.0 (19.0, 58.0) | 79.0 (77.0, 84.0) | 84.0 (61.0, 95.0) | 79.0 (19.0, 95.0) |
| **Sex** |  |  |  |  |
| M | 10 (66.7%) | 9 (56.3%) | 7 (53.8%) | 26 (59.1%) |
| F | 5 (33.3%) | 7 (43.8%) | 6 (46.2%) | 18 (40.9%) |
| **Brain pH** |  |  |  |  |
| Mean (SD) | 6.26 (0.218) | 6.05 (0.188) | 6.12 (0.186) | 6.15 (0.213) |
| Median (Min, Max) | 6.27 (5.69, 6.73) | 6.01 (5.76, 6.50) | 6.10 (5.82, 6.55) | 6.13 (5.69, 6.73) |
| **PMI (Hours)** |  |  |  |  |
| Mean (SD) | 86.7 (22.9) | 61.5 (18.4) | 83.4 (17.8) | 76.5 (22.6) |
| Median (Min, Max) | 94.0 (47.0, 126) | 64.5 (30.0, 95.0) | 80.0 (49.0, 112) | 74.5 (30.0, 126) |
| **Brain Weight (g)** |  |  |  |  |
| Mean (SD) | 1430 (111) | 1330 (99.2) | 1240 (134) | 1330 (136) |
| Median (Min, Max) | 1460 (1200, 1570) | 1320 (1160, 1500) | 1220 (1030, 1440) | 1320 (1030, 1570) |
| **APOE Genotype\*\*** |  |  |  |  |
| 2/3 | 0 (0%) | 1 (6.3%) | 0 (0%) | 1 (2.3%) |
| 3/3 | 9 (60.0%) | 12 (75.0%) | 3 (23.1%) | 24 (54.5%) |
| 3/4 | 6 (40.0%) | 3 (18.8%) | 10 (76.9%) | 19 (43.2%) |
| **Braak Stage** |  |  |  |  |
| 0 | 15 (100%) | 2 (12.5%) | 0 (0%) | 2 (4.5%) |
| 1 | 0 (0%) | 6 (37.5%) | 0 (0%) | 6 (13.6%) |
| 2 | 0 (0%) | 4 (25.0%) | 0 (0%) | 4 (9.1%) |
| 3 | 0 (0%) | 1 (6.2%) | 0 (0%) | 1 (2.3%) |
| 4 | 0 (0%) | 2 (12.5%) | 0 (0%) | 2 (4.5%) |
| 5 | 0 (0%) | 0 (0%) | 0 (0%) | 0 (0%) |
| 6 | 0 (0%) | 1 (6.2%) | 13 (100%) | 14 (31.8%) |
| **Thal Stage** |  |  |  |  |
| 0 | 13 (86.7%) | 3 (18.8%) | 0 (0%) | 3 (6.8%) |
| 1 | 2 (13.3%) | 6 (37.5%) | 0 (0%) | 6 (13.6%) |
| 2 | 0 (0%) | 2 (12.5%) | 0 (0%) | 2 (4.5%) |
| 3 | 0 (0%) | 2 (12.5%) | 1 (7.7%) | 3 (6.8%) |
| 4 | 0 (0%) | 2 (12.5%) | 2 (15.4%) | 4 (9.1%) |
| 5 | 0 (0%) | 1 (6.3%) | 10 (76.9%) | 11 (25.0%) |
| 6 | 0 (0%) | 0 (0%) | 0 (0%) | 0 (0%) |

\*HA subset of LBC1936 Cohort
\*\*No 2/4 or 4/4 APOE genotypes

### Cognitive testing

The Moray House Test (MHT) of general intelligence was administered to LBC1936 participants at age 11 and at ages 70, 76, and 79. Longitudinal MHT scores were used to sub-categorise donors of the available post-mortem brain tissue samples from our healthy agers (HA) into either Lifetime cognitive resilient (LCR) or Lifetime cognitive decline (LCD) groups. This classification was achieved by plotting age-adjusted MHT scores from age 11 against the mean of older-age-adjusted MHT scores from ages 70 and 76 from all healthy agers in the cohort (n = 641). Donors of post-mortem samples were then plotted on this lifetime cognitive scale, where samples above the regression line were defined as LCR (n= 8) and those below were defined as LCD (n=7).

### Tissue preparation

Protocols for post-mortem brain processing were detailed previously [23]. Brain tissues were processed for biochemistry and synaptoneurosome preparation and embedding for histopathology (IHC) and array tomography (AT) (Supplementary Methods Fig. 2). Brain regions studied included: primary visual cortex (BA17); middle temporal gyrus (BA20/21); anterior cingulate cortex (BA24); dorsolateral prefrontal cortex (BA46) and posterior hippocampus. For AT, RNA-seq and proteomics, two of these brain regions BA17 and BA20/21 were investigated.

### Synaptoneurosome preparation

Total tissue homogenate and synaptoneurosome fractions were prepared using frozen tissue samples with minor adjustments as previously described [24]. Tissue was homogenized and passed through an 80μm nylon net filter (Millipore) to generate the total homogenate. Half of the total homogenate was passed through a Millex-SV 5μm membrane filter (Millipore) then centrifuged at 1000 × g for 5 minutes to generate synaptoneurosome pellets. Pellets were reconstituted and homogenised with 450μL buffer. Protein quantification was carried out using the micro Bicinchoninic acid assay (Pierce, UK).

### Label-Free quantitative (LFQ) mass spectrometry (MS)

Sample preparation for LFQ-MS was carried out based on previous proteomic workflows [25, 26]. Peptides generated from total homogenate and synaptoneurosome samples were analysed by MS using a Q-Exactive-HF (Thermo Scientific) mass spectrometer coupled with a dionex Ultimate 3000 RSLCnano (Thermo Scientific). Protein differential expression analysis was performed using DEP (Differential Enrichment analysis of Proteomics data; R package version 1.6.1).

### SDS-PAGE and immunoblotting

SDS-PAGE and immunoblotting was performed as described previously [23]. Primary antibodies used for immunoblotting are shown in Supplementary methods (Supplementary Methods Table 3). Proteins were detected on an Odyssey system using 680 and 800 IR dye secondary antibodies. Total protein stains were performed with Ponceau S and/or REVERT total protein stains.

### RNA extraction and analysis

mRNA was extracted from both total homogenate and synaptoneurosome samples using the RNeasy Plus Micro Kit (Qiagen). Illumina libraries were prepared using TruSeq mRNA Sample Prep Kit. Illumina sequencing was carried out on a NovaSeq platform using 50 base paired-end reads. Differential expression analysis was performed using DESeq2 (R package version 1.24.0) [27] followed by gene set analysis using Camera [28] from the limma R package (version 3.40.6) [29] and Gene Ontology enrichment analysis using topGO (R package version 2.36; (https://rdrr.io/bioc/topGO/). qPCR was performed in an CFX96 Real-time system (Bio-Rad) using BRYT Green Dye (Promega) and the GoTaq 1-Step RT-qPCR kit (Promega).

### Histopathology

Fresh post-mortem tissue blocks were fixed in 10% formalin, dehydrated and paraffin embedded. Tissue sections were cut on a Leica microtome at 4μm thickness and processed for immunohistochemistry using the Novolink Polymer Detection Kit (Leica, Supplementary Fig. 1). All immunolabelled sections were assessed blind to case information and stain burdens calculated using Stereo Investigator (MBF Bioscience).

### Array Tomography

Fresh brain samples were fixed in 4% paraformaldehyde, dehydrated, and embedded in LR white resin as previously described [22]. Ribbons of 70nm serial sections were cut on an ultramicrotome (Leica) then stained with immunofluorescence and imaged using an AxioImager Z2 with an 63x 1.4 NA objective. Array tomography images were processed with an in-house image analysis pipeline (Supplementary Fig. 1).

### iPSC-neuron culture

We reprogrammed peripheral blood mononuclear cells (PBMCs) from the LBC1936 cohort as detailed in [30]. iPSCs were differentiated to glutamatergic cortical neurons by dual SMAD inhibition, following a protocol adapted from Shi et al. [31]. Neurons were cultured to 60 days post final passage *in vitro*, exposed to Aβ+ or Aβ-homogenate for 48 hours, then samples collected for RNA extraction or fixed for imaging. Stained neurons were imaged on a Leica TCS confocal microscope with an oil immersion 63x objective. Generation and immunodepletion of human brain homogenate for treating iPSC derived neurons was conducted following a protocol adapted from Hong et al. [32]. Human superior temporal cortex was homogenised, centrifuged, and dialyzed. Homogenates were immunodepleted for Aβ using 4G8 antibody or mock immunodepleted with mouse serum. Concentration of Aβ1-42 in Aβ+ and Aβ-homogenate was quantified by ELISA (WAKO, 296-64401).

### Statistics and data sharing

Group comparisons including variables Cohort (or cognitive status), Sex, PMI, APOE status, Plaque present/absent) were analysed using Linear mixed effects models including case as a random effect to account for multiple measures per case. Specifically for array tomography, tissue sample was nested within case as a random effect and experimenter was included as a random effect as more than one person collected the data. Where data failed to meet model assumptions, data were transformed via Tukey’s Ladder of Power. ANOVA with Satterthwaite correction was performed on the linear mixed effects models and post-hoc Tukey-corrected comparisons were made between groups. All analyses were performed using R Studio [33] (R 4.4.1). All software macros, R scripts, and analyzed data spreadsheets containing anonymized data are freely available on Edinburgh DataShare and GitHub.

## RESULTS

### Synaptic pathology and gliosis increase from mid-life to ageing to Alzheimer’s disease

Synapses from two brain regions BA20/21 and BA17 (middle/inferior temporal gyrus and primary visual cortex) were stained for presynapses (synaptophysin), postsynapses (PSD95), tau (total tau) and oligomeric Aβ (OC, Fig. 1). A mean of 341 million paired synapses (SD 53.8 million) were analysed per cohort. There is a stepwise decrease in synapse density from midlife to healthy ageing to AD (post/pre-synaptic pairs defined as a positive pre and post-synaptic puncta within 0.5μm distance) (Fig. 1B). Three-dimensional reconstructions of array tomography image stacks show accumulation of Aβ (Fig. 1C) and tau (Fig. 1D) within synapses. Quantification of co-localization reveals that Aβ accumulates within paired pre-synaptic terminals increasing from mid-life to healthy ageing to AD. Cohort and presence of plaque in the image stack were associated with a significant increase in the proportion of synapses with Aβ (Fig. 1E). Similar results are observed when we examine post-synaptic terminals containing Aβ (Fig. 1F). We observe tau accumulating in pre-synaptic terminals (Fig. 1G) with more tau in AD pre-synapses than in healthy ageing or mid-life, more in BA20/21 than BA17, and more when there is a plaque in the image. Similar effects are observed for tau accumulation in post-synaptic densities (Fig. 1H). Using standard immunohistochemistry on paraffin sections, we measured the accumulation of amyloid plaques and gliosis in five brain regions (BA20/21 -middle/inferior temporal gyrus, BA17 - primary visual cortex, BA24 – cingulate cortex, BA46 – dorsolateral prefrontal cortex, and hippocampus). Aβ accumulation increased between ML, HA and AD post-mortem brains (Supplementary Fig. 2A). Stereological quantification validated a step-wise increase in Aβ between ML, HA and AD samples (Supplementary Fig. 2B) and both cohort and regional effects were evident and highest in BA24 AD brains. Stereological quantification of reactive astrocyte (GFAP) burdens shows an increase between ML, HA and AD (Supplementary Fig. 2C). There was a main effect of region and post-hoc comparisons showed a significant increase in GFAP burden in AD in comparison to HA or ML in BA20/21. Microglial (CD68) burden increased between ML, HA and AD and was different between brain regions with the most pronounced microglial coverage in hippocampus (Supplementary Fig. 2D). Together, these data indicate that synapses are lost between midlife, ageing and AD alongside an increase in gliosis and synaptic accumulation of pathological proteins.

**Fig. 1.**
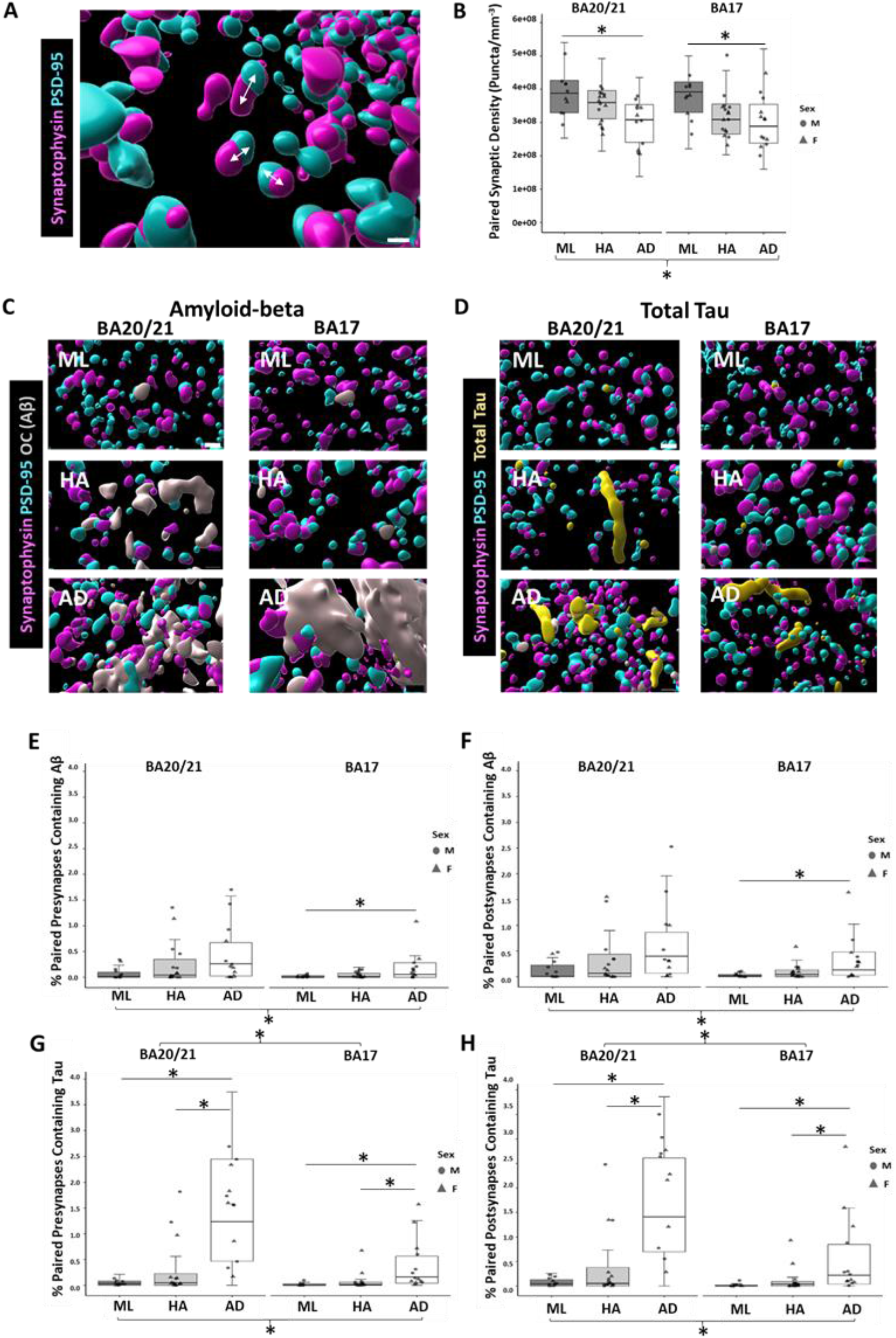
High-resolution array tomography (AT) reveals synaptic pathology in ageing and AD. **A** Synapses were quantified from AT image stacks and counted as a synaptic pair if the centroid of the presynaptic object (synaptophysin, magenta) was within 0.5 mm of the nearest postsynaptic object (PSD95, cyan). Arrows show examples of synaptic pairs. **B** The density of paired synapses decreases between mid life (ML), healthy ageing (HA) and Alzheimer’s disease (AD) and a linear mixed effects model followed by ANOVA shows there was an effect of cohort F[2,33.59]=5.79, p=0.006. There were no differences in synapse density between males and females, or *APOE4* carriers and non-carriers. Pairwise post-hoc comparisons showed synapses were significantly decreased in the AD cohort in comparison to ML and this was evident in both brain regions (BA20/21, t ratio = 3.02; p = 0.01, d = 60; BA17, t ratio = 2.87; p = 0.01, d = 52). We examined the synaptic localisation of Aβ (grey, **C**) and tau (yellow, **D**). In presynaptic terminals, there was a trend towards an increase in Aβ accumulation between midlife, healthy agers, and AD (**E**, F[2,33.82]=3.14, p=0.05) and a significant increase in presynaptic Aβ in regions containing plaques (F[2,157.48]=16.47, p<0.0001). Pairwise post-hoc comparisons showed a significant increase in Aβ accumulation in AD BA17 (t ratio = 2.63; p = 0.02, d = 61). In post-synaptic terminals, there was a significant increase in accumulation of Aβ between ML, HA, and AD (**F**, F[2,35.62]=3.91, p=0.03) and a significant increase in postsynaptic Aβ in regions containing plaques (F[2,150.86]=13.03, p<0.0001). Pairwise post-hoc comparisons showed a significant increase in Aβ accumulation in AD BA17 (t ratio = 2.81; p = 0.01, d = 56). Presynaptic tau accumulation was significantly increased in AD (**G**, F[2,30.28]=26.66, p<0.0001) and significantly higher in BA20/21 than in BA17 (F[1,134.08]=27.23, p<0.001). There was also a significant interaction between cohort and brain region (F[2,126.09]=4.15, p=0.02) and a trend toward increased presynaptic tau in regions containing amyloid plaques (F[2,144.71]=2.56, p=0.08). Pairwise post-hoc comparisons showed a significant increase in presynaptic tau accumulation in AD in comparison to ML and HA cohorts and this was evident in both regions (BA20/21, ML v AD (t ratio = 5.91; p = <0.0001, d = 60); HA v AD (t ratio = 6.57; p = <0.0001, d = 48); BA17, ML v AD (t ratio = 4.29; p = 0.002, d = 52); HA v AD (t ratio = 4.32; p = 0.002, d = 46). Postsynaptic tau accumulation increases from ML to HA to AD groups (**H**, F[2,30.35]=29.54, p<0.0001) and is higher in BA20/21 than BA17 (F[1,134.6]=34.2, p<0.001). There is also an interaction between brain region and cohort (F[2,126.15]=5.72, p=0.004). Pairwise post-hoc comparisons showed a significant increase in postsynaptic tau accumulation in AD in comparison to ML and HA cohorts and this was evident in both regions (BA20/21, ML v AD (t ratio = 6.17; p = <0.0001, d = 60); HA v AD (t ratio = 7.15; p = <0.0001, d = 48); BA17, ML v AD (t ratio = 4.46; p = 0.0001, d = 47); HA v AD (t ratio = 4.47; p = 0.0001, d = 47). For box-plots, each point represents case medians. Statistical analyses carried out using linear mixed effects models followed by ANOVA and post-hoc comparisons. Scale bar 1μm for IMARIS reconstructions.

**Fig. 2.**
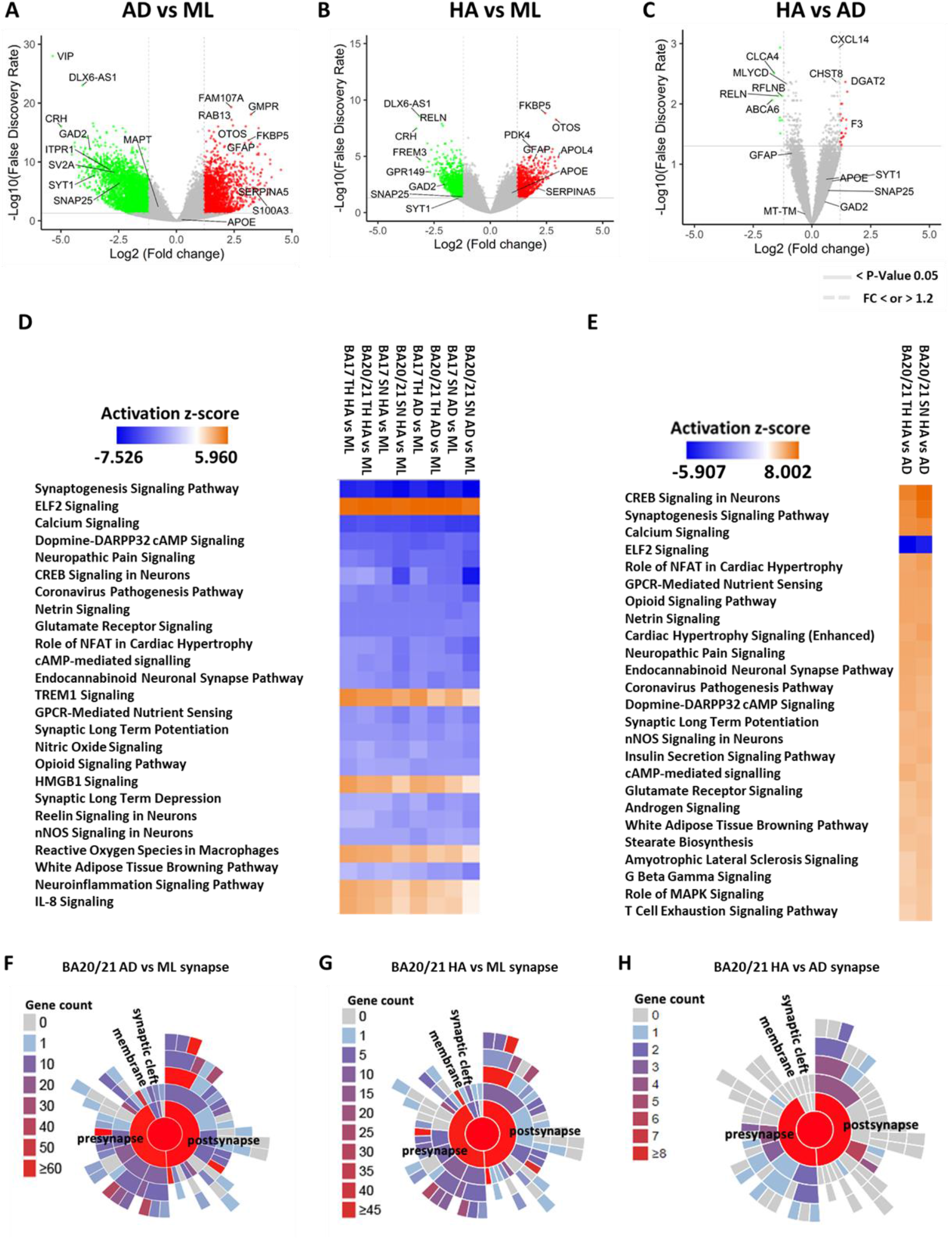
Molecular changes in synapses in healthy ageing (HA) and AD indicate decreased synaptic function and increased inflammation compared to mid-life (ML). **A** Comparing BA20/21 synaptic (synaptoneurosome) transcriptional changes between AD versus ML highlights many differentially expressed genes (9,671 DEG’s < FDR 0.05). **B** 5,801 DEGs with FDR<0.05 between HA and ML. **C** Fewer transcriptional changes are observed between HA and AD (293 DEG’s < FDR 0.05). **D** Top 25 canonical pathways associated with total homogenate (TH) and synaptoneurosome (SN) brain preparations across both brain regions (BA17 and BA20/21) display similar profiles between HA or AD to ML cohorts. An inhibition or a decrease of canonical pathways (blue) associated with neurotransmission and memory is evident in both HA and AD in comparison to ML cohorts. Stress and immune response pathways appear to be activated (orange) or increased in both HA and AD in comparison to ML. **E** The top 25 canonical pathways associated with TH and SN brain preparations in BA20/21 between HA versus AD (BA17 showed no differences) show a decrease in only one biological pathway (blue) associated with stress response, whilst remaining canonical pathways associated with neurotransmission and memory were all increased (orange) in HA cohort. **F** 894 of the DEGs between AD and ML cohorts in BA20/21 synapses mapped to known synaptic proteins in the SynGO annotated database. 467 of these DEGs were associated with the post-synapse and 401 with the pre-synapse. **G** In HA versus ML BA20/21 synapses, 615 genes were mapped to SynGO of which 337 were associated with the post-synapse and 285 with the pre-synapse. **H** From the 293 transcripts identified in synapses of BA20/21 (HA vs AD), an even number of both pre and post-synaptic genes (n = 24) were identified using SynGo curated database, highlighting both synapse domains were adapting equalling in the healthy agers and/or not adapting in the AD brains. **A-C** show Volcano plots of log2 fold change vs -log10 of the false discovery rate. Genes above solid grey line on volcano plots show FDR=0.05 and dotted lines log2 fold change 1.2 (red) and -1.2 (green) respectively. Transcripts of interest are labelled in black.

### Molecular changes in synapses in age and Alzheimer’s disease

Using RNA sequencing, we observe thousands of differentially expressed gene transcripts (DEG’s) in synapses from BA20/21 temporal cortex samples from people with AD compared to mid-life controls (9,671 DEG’s) and when comparing healthy ageing people without dementia (HA) to mid-life controls (5,801 DEG’s, Fig. 2A-B). 293 genes were differentially expressed between HA and AD subjects (Fig. 2C). Similarly, numerous transcriptional changes were also observed in BA17 region and in total homogenate preparations between AD/HA and ML, but there were no differences in gene expression in BA17 between healthy ageing and AD (DOI Supplementary Table 2). Canonical pathways predicted to be inhibited (decreased) in AD or HA in comparison to ML were associated with neurotransmission and memory whilst stress and immune response pathways were predicated to be activated or increased (Fig. 2D). In HA vs AD, there were increases in synaptic function pathways and a decrease in one stress response pathway, EIF2 signalling (Fig. 2E). These data compliment Gene-set analysis (GSA) datasets (DOI Supplementary Folder 1-2) generated independently of IPA and based on gene sets with shared biological and functional properties. Using the SynGo curated database (https://www.syngoportal.org/index.html) specifically designed as a resource for synaptic specific associated genes, we observe approximately equal numbers of changes in genes important in pre and post synaptic function (Fig. 2F-H). Fewer differentially expressed proteins (DEP’s) were identified (DOI Supplementary Table 2) when comparing the same tissues at the proteomic level (Supplementary Fig. 3A-C). A subset of DEG’s identified from RNA-seq datasets were validated by RT-qPCR (Supplementary Fig. 3D-J) namely, Related Ras small GTPase (*RRAS)*; Inositol 1,4,5-trisphosphate (*ITPR1*), Glutamate decarboxylase 2 (*GAD2*); Synaptosomal-Associated Protein, 25kDa (*SNAP-25*), Transmembrane 4 L Six Family Member 1 (*TM4SF1*) and Synatotagamin-1 (*SYT1*). Overall, all RT-qPCR expression profiles matched RNA-seq directional changes (Supplementary Table 3). A subset of DEP’s namely, Clusterin (CLU), Stomatin (STOM) and Vimentin (VIM) identified from proteomics datasets were validated by immunoblotting (Supplementary Fig. 3K-N). Collectively, all immunolabelling blot expression profiles matched proteomic directional changes (Supplementary Table 3).

### Cognitive resilience in healthy ageing is associated with lower glial burdens and dampened synaptic activity pathways

In addition to examining differences between healthy ageing, mid-life and Alzheimer’s disease, we stratified healthy ageing participants into categories based on their lifetime cognitive trajectories. To determine cognitive ageing status as either lifetime cognitive resilience (LCR) or lifetime cognitive decline (LCD), we plotted the age adjusted intelligence test scores at age 11 years versus the mean of those taken at age 70 and 76 for all participants in the LBC1936 study with relevant data (Supplementary Methods Fig. 1). Brain donors in this study who fall above the population regression line were considered resilient (n = 8) and those below were categorised as having lifetime cognitive decline (n = 7). Array tomography analysis revealed no differences in synapse densities between cognitive groups however densities were lower in BA17 than in BA20/21 (Fig. 3A, B). These data indicate the possibility that other mediators may be affecting cognitive performance in ageing. Next, we investigated whether Aβ and/or tau co-localization differs in those considered cognitively resilient compared to people falling into the lifetime cognitive decline category. Three-dimensional reconstructions of array tomography image stacks shows no difference in synaptic accumulation of Aβ or tau in LCR vs LCD groups with both cognitive groups displaying more synaptic pathology in BA20/21 than BA17 (Fig. 3C-F). Stereological quantification of Aβ burden across 5 brain regions showed no difference between the cognitive groups but regional variability (Fig. 3G-H). Next, we sought to investigate glia, integral to normal brain function, in relation to lifetime cognitive performances within the HA cohort. Immunohistochemistry imaging shows CD68 burden is increased in the LCD group across all five brain regions (Fig. 3I). The hippocampal region displayed the greatest increase suggesting activated microglia may be a factor in resilience to cognitive ageing. GFAP immunolabelling also shows higher burdens in LCD group across all five brain regions (Fig. 3J).

**Fig. 3.**
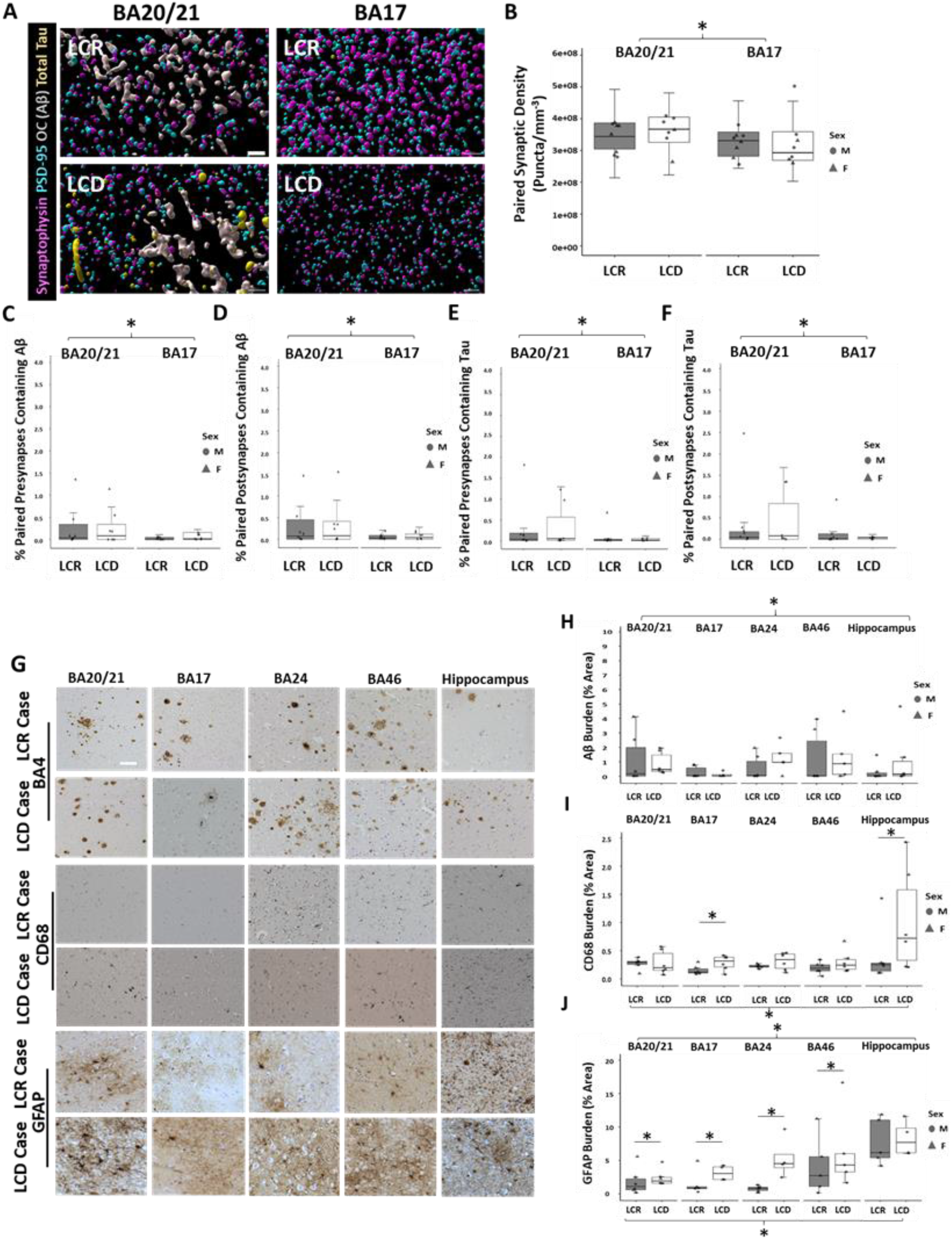
Maintained synaptic density and increased gliosis in people with lifetime cognitive decline. **A** Representative 3D reconstructions of AT stacks from Lifetime cognitive resilient (LCR) and Lifetime cognitive decline (LCD) cohorts. Serial sections of 70 nm sections from BA20/21 and BA17 were stained for synaptophysin (magenta), PSD95 (cyan), OC (grey) and Total tau (yellow). **B** Synapse density was lower in BA17 than BA2021 (F[1,88.93]=4.00, p=0.049); however there was no difference in excitatory synapse density between cognitive cohorts (F[1, 10.67]=0.01, p=0.92). There was similarly no difference between LCR and LCD in the percent pre or post synapses containing Aβ or tau, but co-localization Aβ or tau with synapses showed regional increases in BA20/21 in comparison to BA17 (**C** F[1,102.40]=8.29, p=0.005; **D** F[1,95.52]=4.70, p=0.033; **E** F[1,72.89]=10.23, p=0.002; **F** F[1,70.36]=8.06, p=0.006). Postsynaptic Aβ accumulation was higher in regions containing a plaque (**D** F[2,51.65]=4.39, p=0.017). **G** Representative images of Aβ (BA4), microglia (CD68) and astrocytes (GFAP) are shown from all five brain regions, BA20/21, BA17, BA24, BA46 and hippocampus across LCR and LCD groups. **H** Aβ burden measurements plotted across five brain regions show regional variation in Aβ burdens between both groups (F[4,36.10]=4.71, p=0.004) and an increase in APOE4 carriers (F[1,6.81]=11.85, p=0.011). **I** CD68 burden measurements show a significant increase in microglial burden in the LCD group (F[1,7]=7.52, p=0.029) which reaches post-hoc pairwise significance in BA17 (t ratio = 2.29; p = 0.02, d = 30) and hippocampus (t ratio = 2.89; p = 0.007, d = 30). There is also a significant increase in microglial burden in APOE4 carriers (F[1,7]=6.49, p=0.038). **J** GFAP burden is higher in the LCD group (F[1,6.29]=18.53, p=0.005) and significantly varies across brain regions (F[4,31.93]=6.92, p=0.0004). Post-hoc pairwise significance evident in BA20/21 (t ratio = 2.07; p = 0.04, d = 29); BA17 (t ratio = 2.30; p = 0.02, d = 33); BA24 (t ratio = 4.15; p = 0.0002, d = 33) and BA46 (t ratio = 2.16; p = 0.04, d = 32). For box-plots, each point represents case medians. Statistical analyses carried out using linear mixed effects models followed by ANOVA and post-hoc comparisons. Scale bar 5μm for IMARIS reconstructions, 150μm for IHC images.

RNA sequencing of biochemically isolated synaptoneurosomes reveals 363 synaptic DEG’s (< FDR 0.05) between LCR and LCD cohorts in BA20/21 (Fig. 4A) and 1,116 DEGs in synapses from BA17 (Fig. 4C). These changes were not reflected at the protein level, which showed no DEP’s < FDR 0.05 were identified (DOI Supplementary Table 2). In the total brain homogenate fraction, transcriptional changes were more limited suggesting altered transport of RNA to synapses. (Fig. 4B, D). Only one DEG from total homogenates of BA20/21 was also significantly changed at the protein level, namely acyl-CoA synthetase family member 2 (ACSF2) (FDR < 0.05, DOI Supplementary Table 2). In BA17, similarly only a single DEP, serine and arginine rich splicing factor 7 (SRSF7) was identified (FDR < 0.05, DOI Supplementary Table 2) and expression was increased in the LCR cohort. The top 25 biological pathways identified from transcriptional changes across both brain regions and sample preparations were similar (Fig. 4E). Interestingly, canonical pathways associated with neurotransmission and memory were inhibited or decreased in the LCR cohort, irrespective of region. Activated or increased pathways were limited (orange) and were associated with stress and immune response. These data compliment gene ontology pathways identified which also showed neurotransmission related pathways such as synaptic vesicle exocytosis, post-synaptic activity, neurotransmitter secretion were all down in the LCR cohort (DOI Supplementary Folder 3). Pathways activated or increased were associated with ribosomal and transcriptional processes. The synaptogenesis signaling pathway was ranked highest across all sample and regional cohorts (Fig. 4E) and was decreased in the LCR group. As shown in Supplementary Fig. 4, this pathway is involved many complex biological interactions at both pre and post-synaptic terminals. CDK5 signaling was ranked 25^th^ on the IPA canonical heatmap (Fig. 4E), and as illustrated in Supplementary Fig. 5, this pathway shows Tau is downregulated in the LCR cohort. Regulation of tau through this pathway may be protective here in the LCR cohort. Together these data indicate that despite similar synapse densities and synaptic accumulation of tau and Aβ, levels of transcripts involved in synaptic signalling are decreased in synapses of people who have maintained cognition, indicating that resilience to pathology-induced hyperactivity may be protective against cognitive decline.

**Fig. 4.**
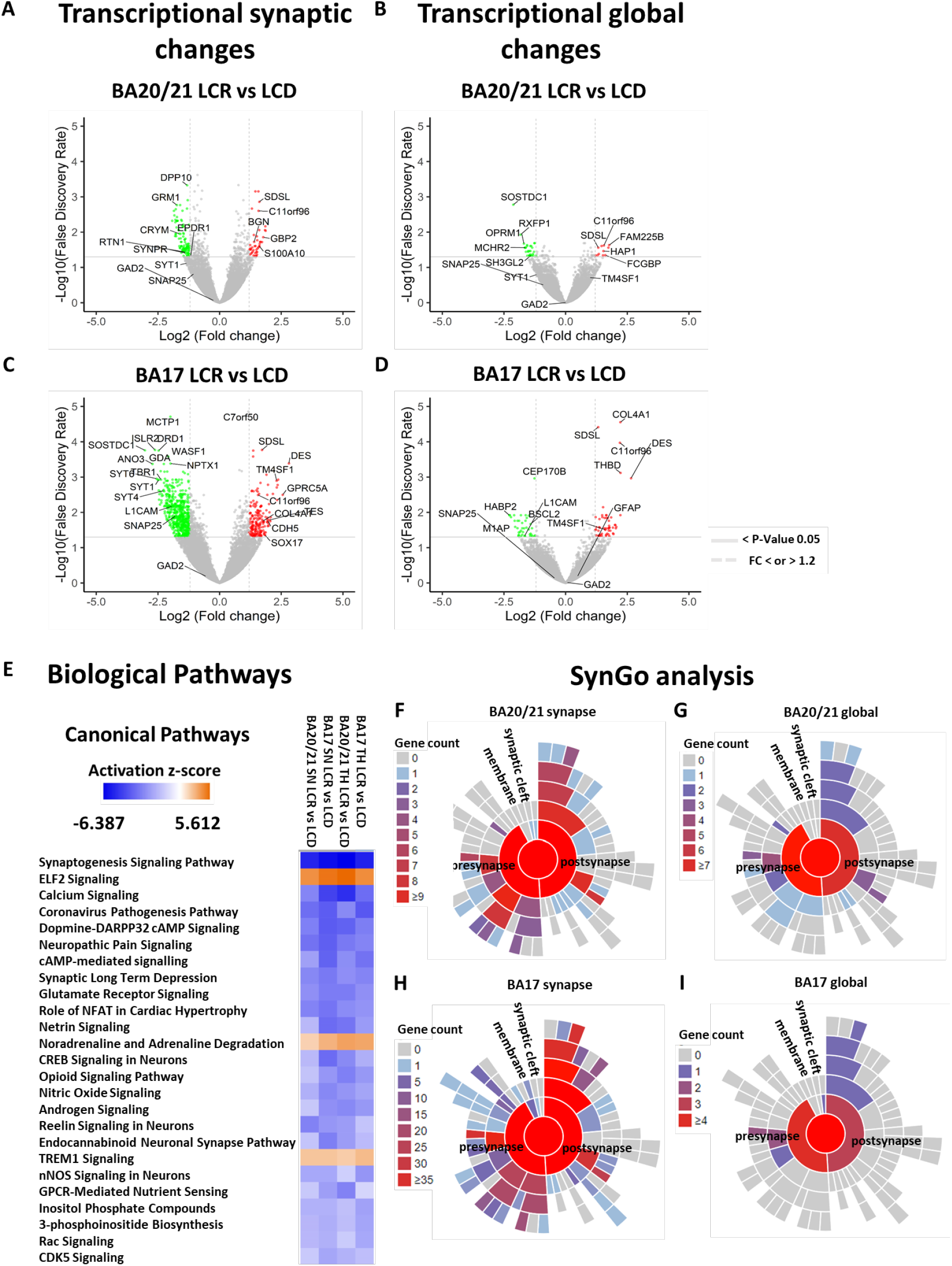
People with lifetime cognitive resilience have dampened synaptic signalling pathways. **A** 363 DEG’s (< FDR 0.05) were found when comparing transcription at the synapse in brain region BA20/21 between LCR and LCD cohorts. **B** Fewer DEG’s (75, < FDR 0.05) were observed in the total homogenate fraction. **C** In BA17, 1,116 DEG’s < FDR 0.05 were identified between cognitive groups. **D** This was not reflected at a global level where only 112 DEG’s were identified. **E** The top 25 canonical pathways changes indicate decreases in abundance (blue) of many pathways involved in synaptic function including neurotransmission and memory in people with lifetime cognitive resilience. Increased pathways (orange) were associated with stress and immune responses. **F** When looking at transcripts in the synapses of BA20/21, there are 363 DEGs of which 27 are known pre-synaptic genes and 27 are known post-synaptic genes in the SynGo curated database. The majority of these (23 of each) are downregulated. **G** SynGo analysis of the 75 DEGs in total homogenate of BA20/21 shows that people with better cognition had alterations in 10 synaptic genes, of which 9 are pre-synaptic and 7 were post-synaptic highlighting overlap between synaptic genes identified. **H** In BA17 synaptic fractions, there are 107 pre-synaptic and 106 post-synaptic genes changed. Of these, the vast majority (103 pre, 102 post) are downregulated. **I** In total 4 pre-synaptic and 3 post-synaptic specific genes were altered in total homogenate of BA17 between the cognitive groups. Volcano plot of log2 fold change vs -log10 of the false discovery rate. Genes above solid grey line on volcano plots show FDR=0.05 and dotted lines log2 fold change 1.2 (red) and -1.2 (green) respectively. Transcripts of interest are labelled in black.

### iPSC-neuron model from LCR and LCD cognitively resilient individuals reveals dampening of synaptic gene expression in response to challenge with human Aβ in lifetime cognitive resisilent neurons compared to neurons from individuals with lifetime cognitive decline

To explore whether dampening of synaptic gene expression in resilient individuals observed post-mortem could be at least in part due to genetic factors, we used iPSC lines generated from blood cells of LBC1936 participants with known cognitive ageing status [30] (Fig. 5A). iPSC-derived cortical neurons were matured for 60 days post-final passage then challenged with human brain homogenate containing soluble Aβ to model exposure of neurons to low levels Aβ during ageing in line with our observations of amyloid deposition in both LCR and LCD brains (Fig. 3). To specifically assess the effect of Aβ on the synaptic structure and neuronal protein expression, we also exposed cultures to the same brain homogenate immunodepleted for Aβ (Aβ-) for 48 hours (Fig. 5B). Quantification of post-synaptic marker homer1 co-localised with dendritic maker MAP2 showed no difference in synaptic density between LCR and LCD lines (LCR Aβ- and Aβ+ four cases; LCD Aβ- and Aβ+ two cases), regardless of treatment condition (Fig. 5C), supporting previous AT findings in PM tissue (Fig. 3B). On a transcriptional level, some genes identified in PM tissue were also present in the iPSC-neuronal model. Interestingly, in LCR iPSC-neurons *SNAP25* and *SYT1* expression decreases in response to Aβ challenge compared to Aβ-challenge and these synaptic genes are also shown to decrease in PM tissue (Fig. 4A-C) in the LCR cohort. The fact that no differences in *TM4SF1* expression were detected (Fig. 5F) shows that not all synaptic genes are necessarily affected by these experimental conditions *in vitro* and therefore that not all of the susceptibility to the response to the ageing brain environment are genetically encoded.

**Fig. 5.**
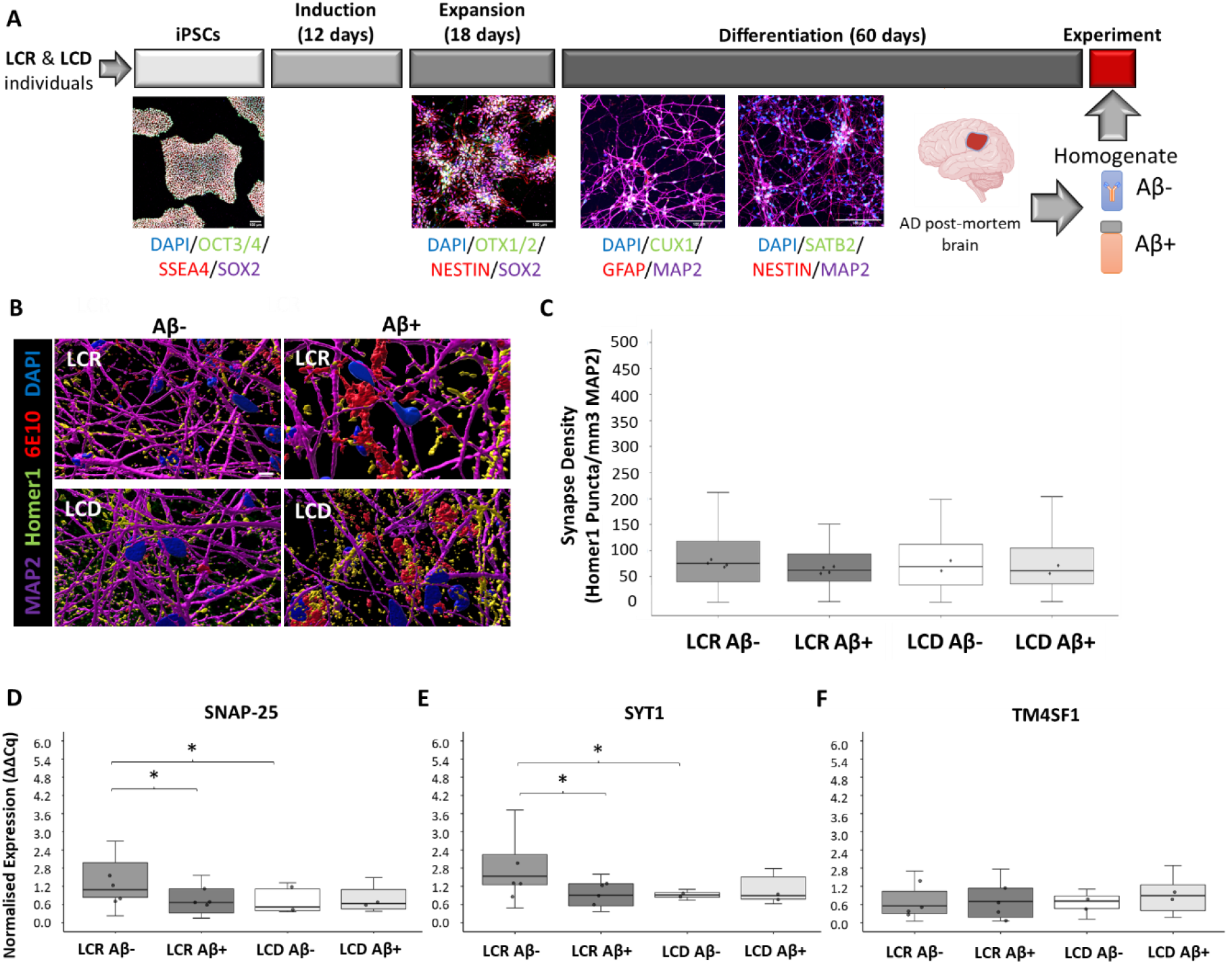
iPSC-derived neurons from LCR individuals have decreased expression of synaptic genes in response to Aβ challenge. **A** Overview of the process by which iPSCs from LCR and LCD individuals were differentiated to cortical neurons. Human AD brain homogenate enriched (Aβ+) or immunodepleted (Aβ-) for soluble Aβ42 was added to cells at experiment time point. **B** Representative 3D reconstructions showing the effect of homogenate treatment on LCD and LCR neurons. **C** Automated quantification of homer1 puncta (post-synaptic density) colocalised with MAP2 (dendrites) showed no significant difference between Aβ treatment or cell line group. **D**-**E** RT-qPCR revealed that *SNAP-25* and *SYT1* expression significantly differed between Aβ treatments in the Lifetime Cognitive Resilient (LCR) group, but not in the Lifetime Cognitive Decline (LCD) group. **F** No difference between treatment or lifetime cognitive ageing groups was observed in *TM4SF1* expression. For box-plots, each point represents case medians. Statistical analysis carried out using linear mixed effects models, synapse density or gene expression ∼ Aβ Treatment * Cognitive Group + (1|Cell Line/Experiment Batch, followed by ANOVA. Scale bar 100 μm (A), 5μm for IMARIS reconstructions (B).

## DISCUSSION

In this study, we present detailed characterisation of post-mortem brain differences in young people, healthy agers and people with AD at the structural and molecular level. We observed a stepwise decrease in paired synaptic densities between young controls (ML), ageing participants with normal cognition (HA), and people with Alzheimer’s disease (AD). The reduction of synapses between HA and ML cohorts was expected as although generally cognitively healthy, HA cases are still within the elderly age range (77-84), and during this normal ageing process a reduction in synaptic numbers of between 10-15% is anticipated after 65 years of age [34]. A stepwise increase in the proportion of remaining synapses containing amyloid beta or tau proteins was also evident, which was more pronounced in BA20/21 than BA17 as expected from the known distribution of pathology in AD. A stepwise increase in GFAP burden between ML, HA and AD was expected as GFAP increases progressively with ageing and cognitive decline [35]. This increase could also be in response to Aβ burden although higher levels of GFAP burdens seen in hippocampus regions across all cohorts do not mirror Aβ burdens in the same region comparison suggesting other factors were at play. Previous studies investigating DNA methylation signatures in the LBC1936 cohort have indicated CD68 burdens are highest in the hippocampal brain region [36]. Here we again show evidence of this neuroimmune response; however, we also highlight significant cohort variation although burden levels were relatively low and below 0.5% for all regions apart from the hippocampus. Collectively, these data highlight the importance of using more than one brain region for investigations as regional differences in burdens can vary, and this may be due to many factors such as cognitive processing demands of that region, pathological load and disease stage.

In addition to pathological studies, we examined RNA and proteins isolated from the same tissue, with the hope of discovering novel biological processes associated with cognitive decline. Our analysis shows limited correlations between both RNA and protein datasets as has been described [37]. One potential explanation for this discrepancy is that large numbers of transcripts are not translated to proteins or are transcribed, but some may be expressed below the detection limit of proteomics. Protein abundance is also controlled by other factors such as post-transcriptional regulation, modifications and degradation, independent to mRNA abundance. Our transcriptomic data were broadly similar to a study comparing AD to control brain tissue [38] (Supplementary Fig. 6); however here we extend to analysis of synaptic transcripts. Our data indicate that molecular pathways important for synapse function decrease from midlife to ageing to AD and conversely that stress and inflammation pathways increase from midlife to ageing to AD. Several genes of interest were identified including *RRAS*, which is involved in axon guidance, angiogenesis and synaptic function and plasticity [39]. *RRAS* was increased in AD in comparison to ML, possibly to protect neurons from neurodegeneration. In contrast, *ITPR1*, a receptor that mediates calcium release from the endoplasmic reticulum [40] was downregulated in AD in comparison to ML. *GAD2* an enzyme involved in the synthesis of the major inhibitory neurotransmitter, γ-aminobutyric acid (GABA) and therefore present in GABAergic inhibitory neurons and synaptic terminals was also downregulated in AD versus ML. These data support our recent findings showing significant decreases in inhibitory neuron and synapse density in AD [41]. Earlier studies in our lab have shown CLU is increased in AD [10] supporting genome wide association studies that previously identified the protein as an additional risk factor involved in AD [42]. Here, we show CLU is upregulated in AD in comparison to ML controls. Interestingly, direct comparison between HA versus AD show no evidence of transcriptional changes in BA17. This implies the BA17 brain region in AD had a similar transcript profile to a healthy aged brain, regardless of dementia status which, supports previous studies showing the visual cortex is affected relatively late in the disease process [43]. Synaptic densities were also in the same range between HA and AD in the BA17 region, supporting this observation. These data again highlight the importance of using more than one brain region for investigations as regional differences in transcriptional expression can vary. The majority of activated canonical pathways in BA20/21 in HA versus AD were associated with neurotransmission and memory, and included biological pathways such as CREB, synaptogenesis, calcium, LTP and glutamate receptor signaling.

Within our HA cohort, age-adjusted longitudinal MHT scores were used to sub-categorise samples into a Lifetime cognitive resilient (LCR) group and a Lifetime cognitive decline (LCD) group. We acknowledge here the further subsetting of these samples into smaller sizes confers risks for unreliable detection of effects due to low statistical power. Nevertheless, life course cognitive data plus post-mortem brain is rare, and characterising changes in brain structure in both groups will further our understanding of healthy brain ageing and cognitive decline. There was no change in synapse density between people with age-related cognitive decline in the absence of dementia in either BA20/21 or BA17. It is possible more subtle differences are occurring that are not detectable by AT. Stereological assessment of CD68 and GFAP burdens between LCR and LCD groups highlight activated microglia and astrocytes increase with poorer cognitive status. In addition to gliosis, we observed molecular changes in synapses between LCR and LCD tissue. Interestingly, in BA17 one of the major biological themes identified from our transcriptomic analysis in IPA was Bruton’s tyrosine kinase (BTK). BTK elevation is found in AD mouse models and human AD brains and is associated with a neuroinflammatory response to extracellular Aβ accumulation and or synaptic loss [98]. BTK is also a known regulator of microglial phagocytosis, and inhibition of BTK signaling is known to decrease microglial uptake of synaptosomes and thus may play a role in maintaining and/or improving cognition [98]. Here, BTK was decreased in the LCR cohort. Therefore, manipulating BTK by using BTK inhibitors may provide a promising strategy moving forward to encourage healthy ageing. Collectively, these data suggest activated phagocytic microglial activity is detrimental to healthy ageing. Biological pathways associated with both regions in LCR and LCD groups indicate damping synapse activity in the more cognitively resilient cohort. We know hyperexcitability is an early signature of neuronal and cognitive dysfunction and studies in animal models have indicated hyperactivity in selective circuits driven by pathologies such as oligomeric Aβ can contribute to cognitive impairment [44]. Both Aβ and tau secretion are also increased with neuronal activity thus dampening activity may essentially slow the accumulation or spread of pathological proteins like soluble oligomers that could drive the gliosis we observe in the LCD group. Modulating this brain hyperexcitability, known to be elevated in people with AD, therefore may provide a novel therapeutic approach to improve cognition and/or long-term cognitive resilience. The current investigation of anti-seizure drugs as treatments for AD [45] may thus also be beneficial in preventing cognitive decline during healthy ageing. These findings could be interpreted to contrast with the idea of cognitive reserve and the protective effect of education against developing dementia. However, it could be the case that enriching activities like education make neural circuits more efficient. An example of this comes from a functional magnetic resonance imaging (fMRI) study measuring the brain activity of professional versus naïve piano players [46]. Here brain activation was found to be lower in the professional pianists suggesting this dampening effect may be beneficial to the brain as energy requirements would be less whilst performing these most complex activities. Our transcriptional data provide a valuable resource of genes to target to support cognitive resilience by lowering synaptic activity. However, one would have to be cautious whilst using drugs to instate these neuronal damping changes and future trials will establish if specific drug treatments are safe, tolerable and effective for memory function stabilisation. Neuromodulation technologies to enhance or suppress activity of the neurons is another promising approach in its infancy with hopes of providing superior therapeutics for many cognitive disorders [47].

This study has limitations. Firstly, synaptoneurosomes (SN) prepared may contain small amounts of non-synaptic neuronal, glial and myelin contaminants. Overall, we tried to take into consideration preserving the balance between the purity of samples and technical feasibility for this study. Secondly, for cognitive classifications, there was variability between the last cognitive test and death, which was several years in some cases. This was recorded in Supplementary Methods Table 1. Thirdly, iPSC-derived neurons represent a developmentally young and reductionist model of tissue complexity, making direct comparison of results with post-mortem tissue difficult. Despite this, synaptic genes relevant to post-mortem data were expressed in iPSC-neurons and form a basis for further exploitation in future experiments. Lastly, we acknowledge sample sizes are low and may be prone to both type 1 and 2 statistical errors; however, the study is a valuable and useful indication of the studies that can be done in future when sample sizes are a magnitude larger. Despite these limitations, the data presented here are a comprehensive examination of synaptic changes in a unique cohort of ageing participants. In summary, we observe that between midlife, healthy ageing, and Alzheimer’s disease, there is substantial gliosis, synapse loss, synaptic accumulation of pathology, and lower levels of transcripts involved in synaptic function. In healthy aged participants without dementia, gliosis was associated with cognitive decline, but in contrast to the AD data, lower levels of gene expression associated with synaptic signalling were paradoxically associated with resilience to cognitive decline. This surprising result has implications for preventing cognitive decline in ageing that will be different from treating people with AD. Together our data indicate that synaptic resilience plays an important role in maintaining cognition during ageing.

## Data Availability

All software macros, R scripts, and analyzed data spreadsheets containing anonymized data are freely available on Edinburgh DataShare and GitHub.

## Acknowledgements

The authors thank all LBC1936 study participants and research team members who have contributed, and continue to contribute, to ongoing studies. LBC1936 is supported by the Biotechnology and Biological Sciences Research Council, and the Economic and Social Research Council [BB/W008793/1] (which supports SEH), Age UK (Disconnected Mind project), the Medical Research Council (MR/M01311/1), and the University of Edinburgh. KH is funded by the Wellome Trust (628TRN R46470). SRC is supported by a Sir Henry Dale Fellowship jointly funded by the Wellcome Trust and the Royal Society (221890/Z/20/Z). JAO is supported by an Economic and Social Research Council new investigator grant (ES/S015604/1); TSJ is supported by the European Research Council (ERC) under the European Union’s Horizon 2020 research and innovation programme (grant agreement no. 681181) and the UK Dementia Research Institute which receives its funding from DRI Ltd, funded by the UK Medical Research Council, Alzheimer’s Society, and Alzheimer’s Research UK. Funding for this study was received from an anonymous industry partner who had no influence over the publication.

