## Supplementary figures for "Synaptic resilience is associated with maintained cognition during ageing"

#### King *et al.* Supplementary Results Figures

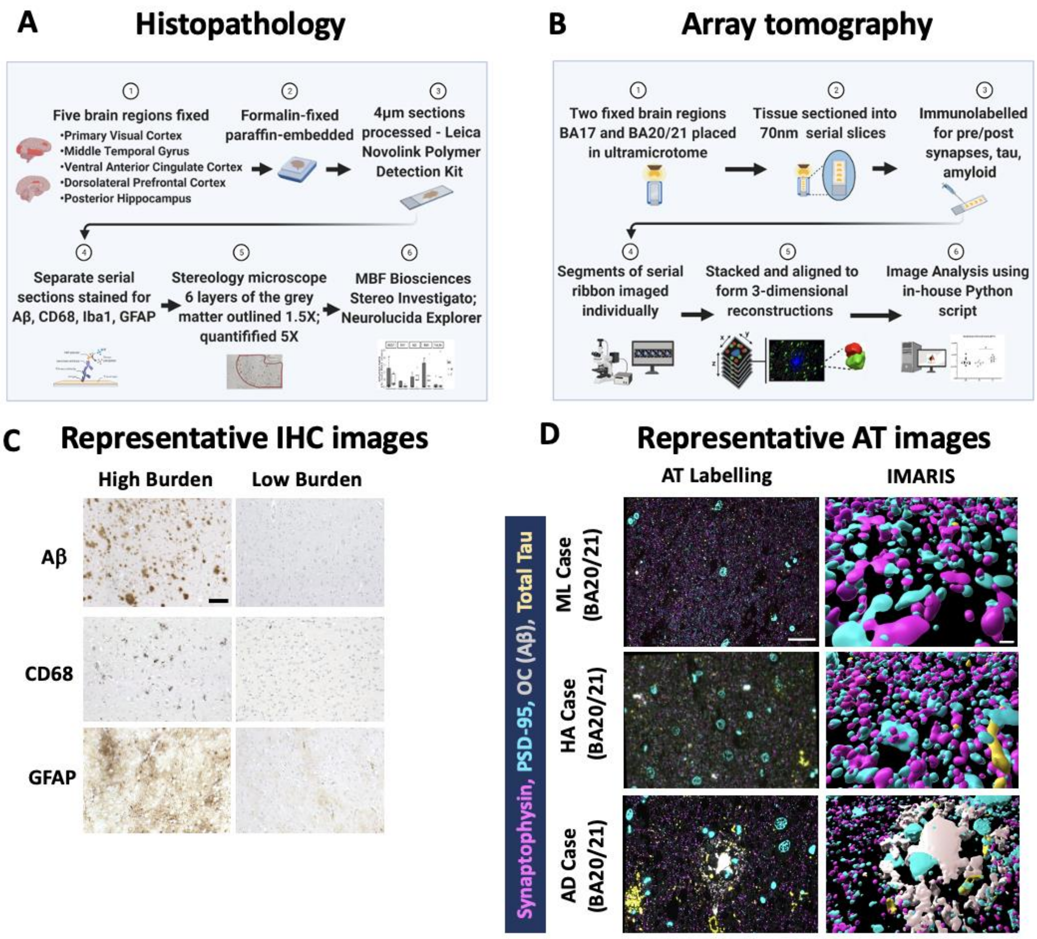


**Supplementary Fig. 1:** Workflows for histopathology (**A**) and array tomography (**B**). Representative IHC images show cases with high vs low burden **(C)**. Array tomography representative images show single section ray images and three-dimensional Imaris reconstructions **(D)**. Scale bars 300µm IHC, 20µm AT immunolabelling and 2µm for Imaris reconstructions.

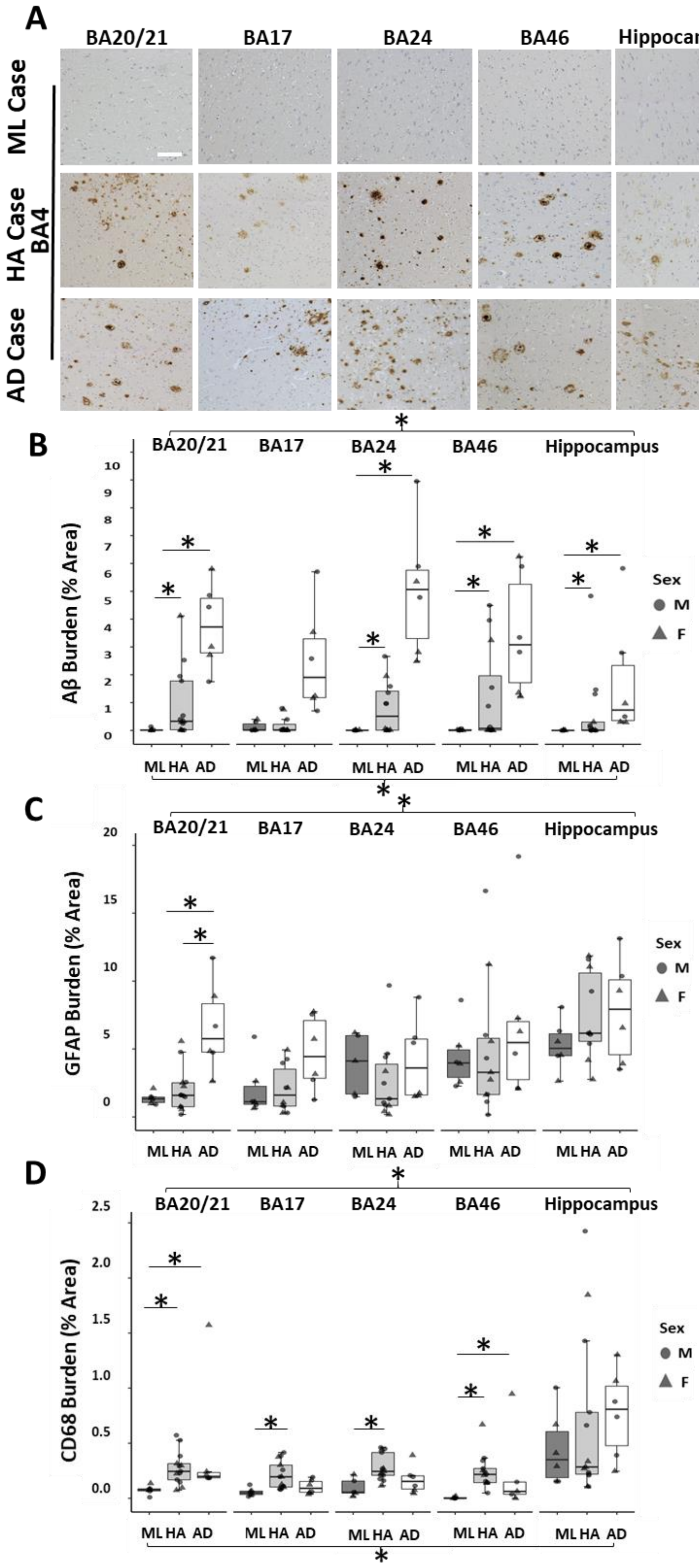
A
BA20/21
BA17
BA24
BA46
Hippocampus
ML Case
HA Case
BA4
AD Case
B
Aβ Burden (% Area)
Sex
M
F
ML HA AD
C
GFAP Burden (% Area)
D
CD68 Burden (% Area)

**Supplementary Fig. 2: Stereological quantification reveals amyloid pathology astrogliosis, and microgliosis from ML to HA to AD.** Amyloid accumulation (**A,B**) was highest in AD cases and varied widely by brain region. Indeed, there was a significant main effect of cohort ($F[2,18.85]=15.85$, $p<0.0001$), region ($F[4, 84.06] = 10.81$, $p <0.0001$) and an interaction effect (Cohort:Region $F[8, 84.08] = 3.30$, $p = 0.002$). APOE4 and male sex were associated with higher amyloid burdens (APOE $F[1, 18.94] = 14.85$, $p = 0.001$; Sex $F[1, 19.19] = 9.2$, $p = 0.007$). GFAP staining was used to measure reactive astrogliosis (**C**). There was a main effect of region $F[4, 78.63] = 8.33$, $p<0.0001$, and post-hoc comparisons showed a significant increase in GFAP burden in AD in comparison to HA in BA20/21. Microgliosis was calculated as CD68 burden (**D**). There was a significant effect of cohort $F[2, 19.44] = 9.38$, $p = 0.001$, region $F[4, 86.65] = 25.84$, $p<0.0001$, and an interaction between Cohort:Region $F[8, 86.60] = 3.73$, $p = 0.0009$. Post-hoc comparisons confirmed microglial activation was significantly lower when comparing ML to either HA or AD across all brain regions. Statistical analyses with linear mixed effects model (Burden ~ Cohort * Region + APOE + Sex + Age_Years + PMI_Hours + (1|Case_BBN), followed by ANOVA. * represent $p<0.05$ Tukey corrected post-hoc comparisons. Scale bar 150 μm.

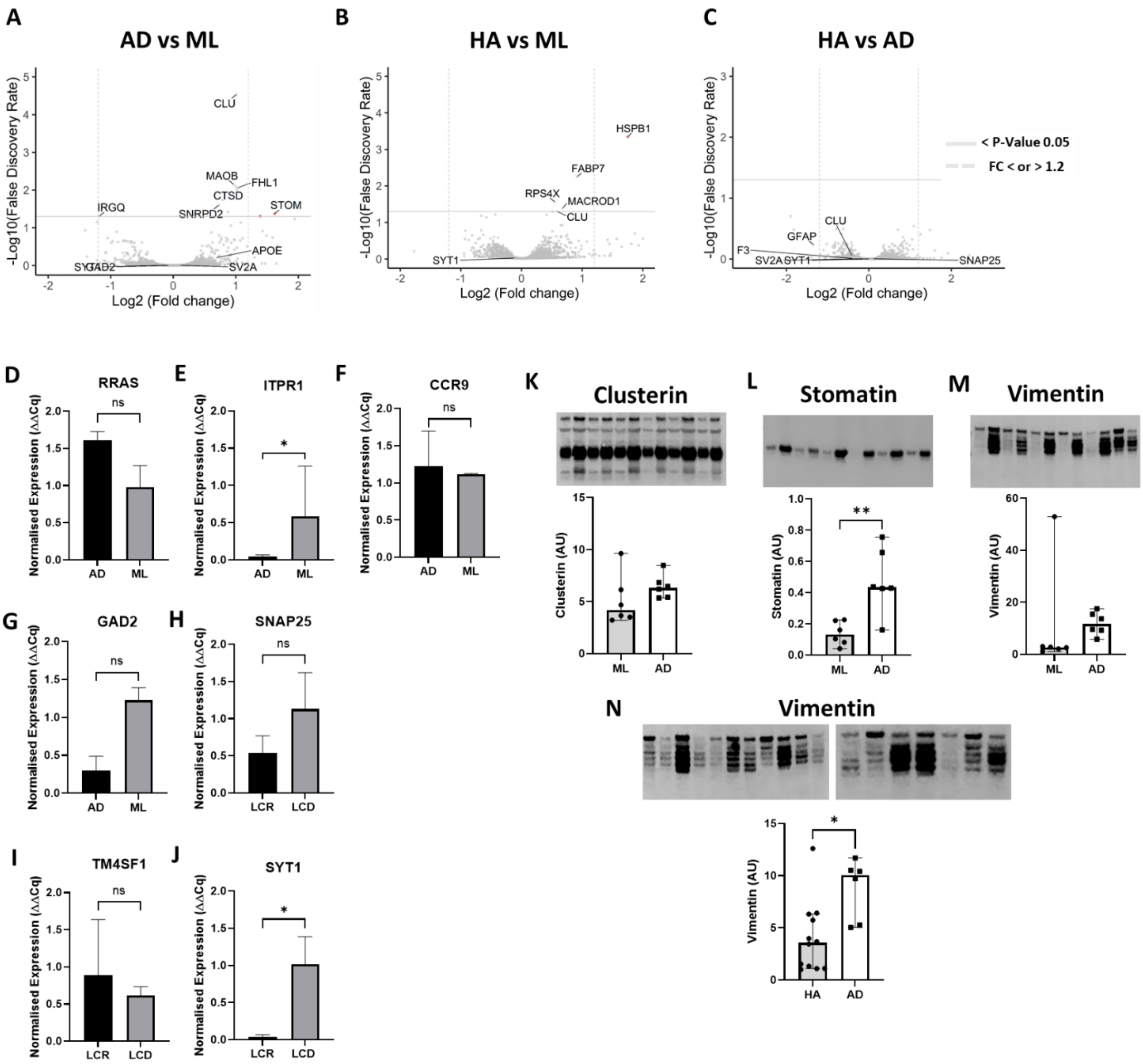

A
AD vs ML
-Log10(False Discovery Rate)
Log2 (Fold change)
CLU
MAOB
FHL1
CTSD
SNRPD2
IRGQ
STOM
APOE
SYGAD2
SV2A
B
HA vs ML
HSPB1
FABP7
RPS4X
MACROD1
CLU
SYT1
C
HA vs AD
< P-Value 0.05
FC < or > 1.2
CLU
GFAP
F3
SV2A SYT1
SNAP25
D
RRAS
E
ITPR1
F
CCR9
G
GAD2
H
SNAP25
I
TM4SF1
J
SYT1
Normalised Expression (ΔΔCq)
ns
*
AD
ML
LCR
LCD
K
Clusterin
Clusterin (AU)
L
Stomatin
Stomatin (AU)
**
M
Vimentin
Vimentin (AU)
N
Vimentin
HA
AD

**Supplementary Fig. 3: Proteomic analysis revels few differentially expressed proteins (DEPs) between cohorts**.

Proteomic DEP changes were more reserved than transcriptomics. **(A)** For AD vs ML only 16 DEP’s (<FDR 0.05) were identified. (**B)** Translational change between HA and ML was also limited (6 DEP’s <FDR 0.05). For HA vs AD, translational DEP identified zero changes (0 DEP’s <FDR 0.05). Volcano plot of log2 fold change vs -log10 of the false discovery rate. Proteins above solid grey line on volcano plots show FDR=0.05 and dotted lines log2 fold change 1.2 (red) and -1.2 (green) respectively. Proteins of interest are labelled in black. Quantitative PCR validation of a subset of RNAseq results **(D-J)**. Differentially expressed genes of interest at the synaptic level (synaptoneurosome) were validated by RT-qPCR. RNA-seq directional observation profiles were confirmed in all cases. RT-qPCR reference genes *CCR9, KCP* and *EN2*, fold changes from RT-qPCR were calculated using 2-ΔΔCt method. Unpaired t tests were carried out in all cases, **D-J**, P-Values 0.3826; 0.402; 0.2808; 0.1208; 0.074; 0.2998 and 0.0291, **D-H**, graphs plotted as median with 95% CI, n=3, n=individual samples, **I-J** mean with 95% CI, n=11. Western blot validation of a subset of proteomics results **(K-N).** Differentially expressed proteins of interest at both global (THp) and synaptic (SNp) levels were validated by western blotting. Proteomic directional observation profiles were confirmed in all cases. **K-L** BA20/21 Synaptoneurosome samples; **M-N** BA20/21 total homogenate samples. REVERT total protein was used in all cases for normalization of target proteins. Unpaired t tests were carried out in all cases, **K-L**, P-Values 0.2599; 0.0038; 0.0649 and 0.0245, graphs plotted as median with 95% CI. **K-M** n=6, **N** n= HA 12, AD n=6, n=individual samples.

### Synaptogenesis Signaling pathway
#### LCR vs LCD

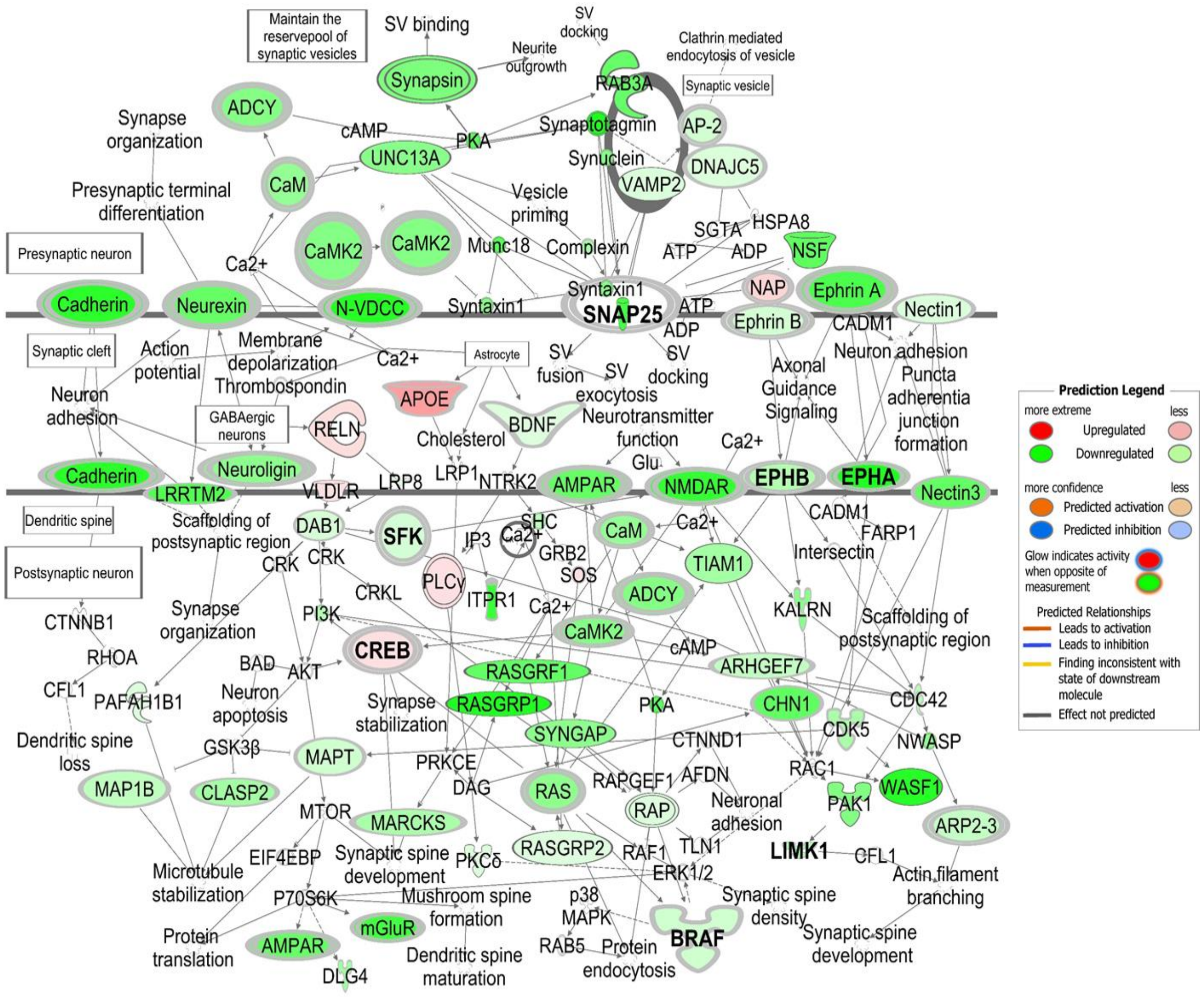


**Supplementary Fig. 4. IPA identified synaptogenesis signaling pathway as top-ranking canonical pathway to be dampened when comparing LCR and LCD samples.** Here we show the pathway specific to BA17 synaptoneurosome LCR vs LCR, however the same pathway applies to BA20/21. Complex biological interactions are shown at the pre-synaptic terminal (*Synapsin*, *SNAP25*); synaptic cleft (*APOE, BDNF*) and at the post-synapse (*MAPT, SYNGAP*) and include 163 molecules in total. Molecules that can be targeted with drugs are shown in bold. At the pre-synapse level *SNAP25* and cadherin can be targeted using botulinum toxin type A and PCA062. At the synaptic cleft *EPHB* and *EPHA* can be manipulated using bosutinib and imatinib. At the post-synaptic level molecules *SFK, BRAF* and *LIMK1* can be targeted using dabrafenib, trametinib or vemurafenib whilst CCS1477 and PRI-724 can be used to target CREB. Collectively, numerous novel and manipulable therapeutic targets are identified here but highlight the multiple drug treatments may be needed to encourage cognitive preservation.

### CDK5 Signaling pathway
### LCR vs LCD

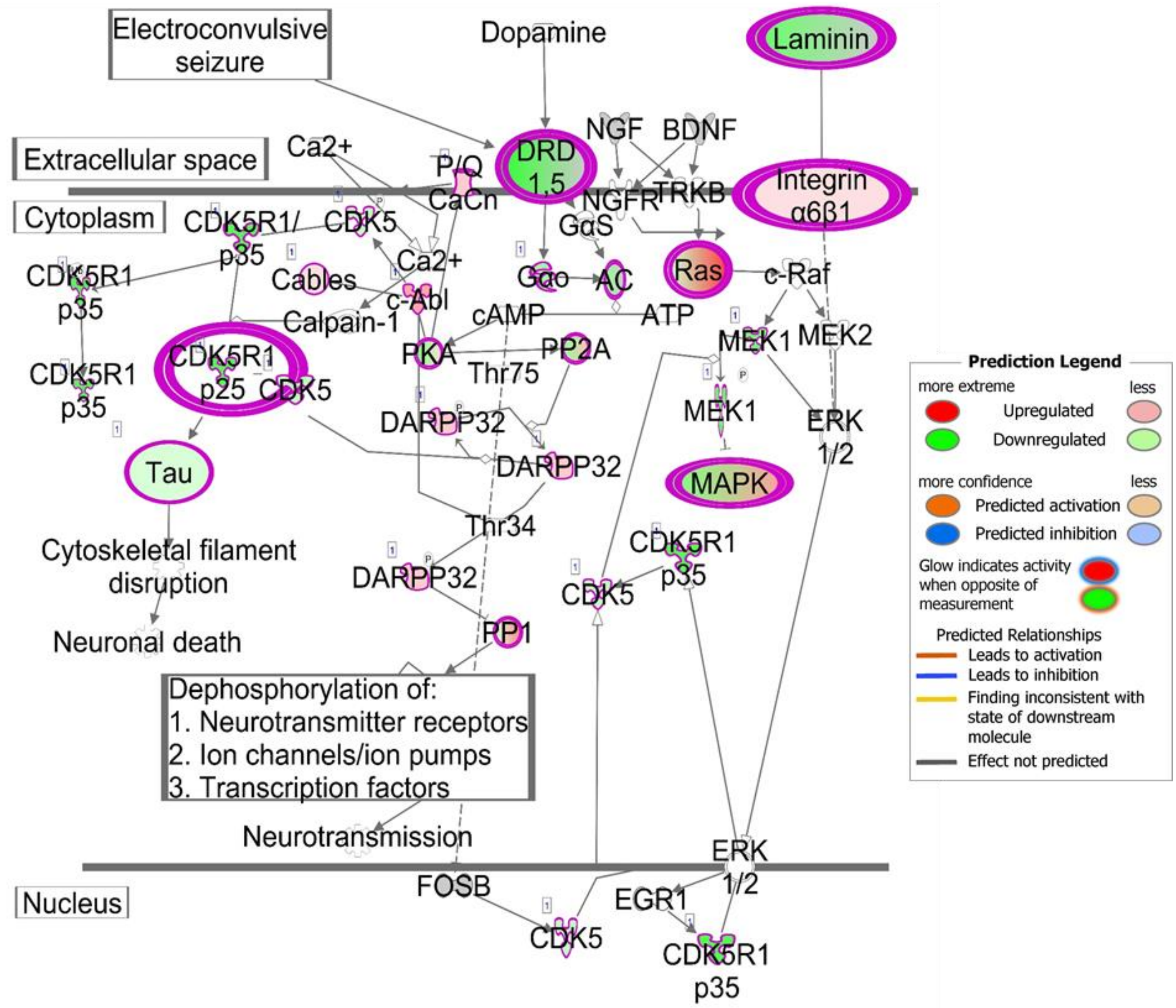

**Supplementary Fig. 5. IPA identified CDK5 signaling pathway as amongst top ranking canonical pathways to change when comparing LCR and LCD samples.** CDK5 signaling pathway was associated with 48 molecules including *Tau* and *Ras* at BA20/21 synapses. *CDK5* substrates include Munc18; Synapsin-1; *PAK1*; Amphiphysin-1; β-Caterin all involved in neuronal migration and neurite outgrowth. Cyclin-dependent kinases (CDKs) are a group of serine/threonine protein kinases which regulate post-mitotic processes such as neuronal activity, neuronal migration during development, and neurite outgrowth. CDK5 also has been implicated in the pathological degeneration of neurons. Indeed, dysregulation of CDK5 can cause the hyperphosphorylation of Tau [83], thereby contributing to neurofibrillary tangle formation, which is the hallmark of AD. It is possible the CDK5R1-p25 complex is inducing a protective effect by downregulating Tau in the LCR cohort and this could be an avenue of focus for future studies.

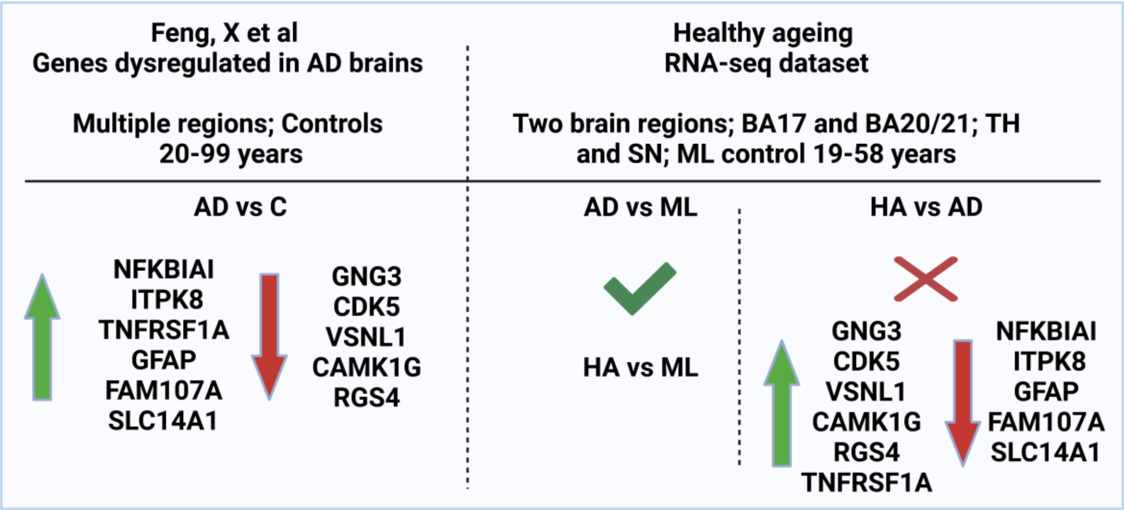


**Supplementary Fig. 6 Transcriptome dataset comparison with AD studies.** A summary of RNA-seq parallels with previous AD transcriptome studies and our healthy ageing dataset. 11 out of 12 genes were present in our heathy ageing dataset for comparison. In both healthy agers (HA) and AD comparisons to control (ML) groups, gene directional changes matched exactly. However, aged, matched controls (healthy agers) compared with AD showed predominantly a reversal in gene directional profiles.
