## supplementary methods for "Synaptic resilience is associated with maintained cognition during ageing"

### King et al Supplementary Methods

***Subjects***

Participants in the study were brain donors to the University of Edinburgh MRC Sudden Death Brain Bank. Donations have been reviewed and approved for use by the Edinburgh Brain Bank ethics committee and the Academic and Clinical Central Office for Research and Development, a joint office of the University of Edinburgh and NHS Lothian (approval 15-HV-016). Three groups of participants were selected: (1) middle aged controls (no known neurological or psychiatric conditions, age range 19-58, n=15); (2) healthy agers from Lothian Birth Cohort 1936 (LBC1936) who did not have dementia or other known neurological or psychiatric conditions (age range 77-84, n=16, note one participant had no available cognitive data) and (3) Alzheimer's disease (with clinical dementia diagnosis and neuropathological diagnosis of AD (age range 61-95, n=13). Samples excluded from healthy agers/LBC1936 groups were re-classified as AD when neuropathology including Braak stage was > 4 and clinical diagnosis was dementia (n = 3). Detailed information for each participant is included in DOI Supplementary Methods Table 1.

***Cognitive testing***

The Moray House Test (MHT) of general intelligence was administered to children at age about 11 years on June 4$^{th}$, 1947. These archived data were followed up decades later when participants were recruited (n = 1,091, LBC1936 cohort) at age about 70 (n=70,805) [1]. Follow-up studies conducted every three (at mean ages of about 73, 76, 79) years thereafter-included detailed cognitive, psychosocial, biomedical, medical, and genetic examinations [2]. MHT tests were included in follow up waves at ages 70, 76, and 79 and were the same MHT test as had been administered at age 11. This well-validated test of general cognitive ability consists of 71 questions (maximum score 76) that involve numerous items such as directions, word classification, reasoning and arithmetic [3] or this study, longitudinal MHT scores were used to sub-categorise donors of the available post-mortem brain tissue samples from our healthy agers (HA; LBC1936) (n = 15, not 16 as one PM brain had no age 11 MHT score) into either Lifetime cognitive resilient (LCR) or Lifetime cognitive decline (LCD) groups. This classification was achieved by plotting age-adjusted MHT scores from age 11 against the mean of older-age-adjusted MHT scores from ages 70 and 76 from all healthy agers in the cohort (n = 641). Donors of post-mortem samples were then plotted on this lifetime cognitive scale, where samples above the regression line were defined as LCR (n= 8), whereas those below were defined as LCD (n=7) (Supplementary Methods Fig. 1). Demographics for cognitive split are shown in Supplementary Table 2 (Supplementary Methods Table 2).

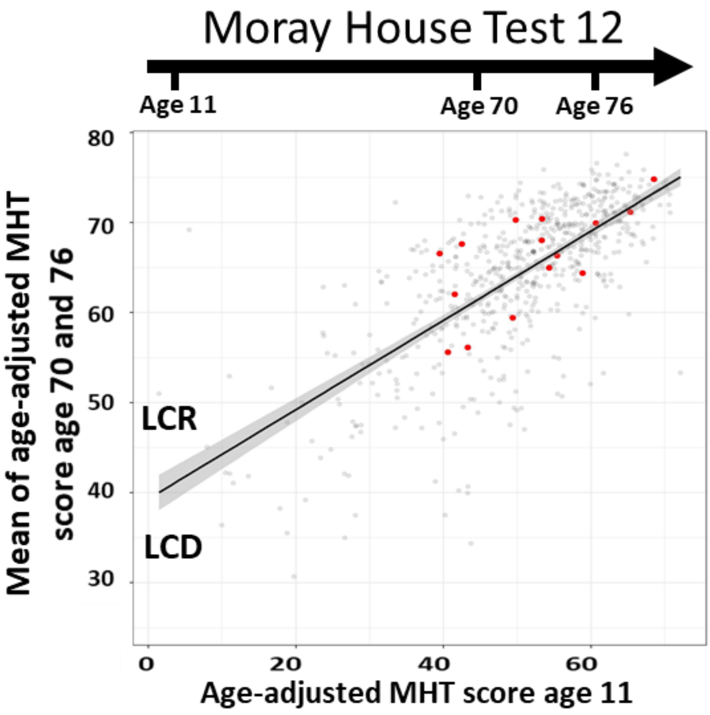


**Supplementary Methods Fig. 1: Longitudinal cognitive ageing.** Age-adjusted longitudinal MHT scores for healthy agers (LBC1936) were plotted as age-adjusted MHT score age 11 (x axis) against mean of age-adjusted MHT score age 70-76 (y axis), n = 641. Available post-mortem samples are shown in red (n = 15). Those above the regression line were categorised as a Lifetime cognitive resilient (LCR) group and those below were labelled as a lifetime cognitive decline (LCD) group. Darker grey shading equals 95% confidence interval.

| Age (Years) | Lifetime Cognitive Resilient (LCR) (N=8) | Lifetime Cognitive Decline (LCD) (N=7) | Overall (N=15) |
|---|---|---|---|
| Mean (SD) | 80.6 (1.92) | 79.9 (2.19) | 80.3 (2.02) |
| Median (Min, Max) | 80.0 (79.0, 84.0) | 79.0 (77.0, 83.0) | 79 (77.0, 84.0) |
| **Sex** | | | |
| M | 4 (50.0%) | 5 (71.4%) | 9 (60.0%) |
| F | 4 (50.0%) | 2 (28.6%) | 6 (40.0%) |
| **Brain pH** | | | |
| Mean (SD) | 5.97 (0.148) | 6.16 (0.196) | 6.06 (0.192) |
| Median (Min, Max) | 5.99 (5.76, 6.20) | 6.13 (5.96. 6.50) | 6.01 (5.76, 6.50) |
| **PMI (Hours)** | | | |
| Mean (SD) | 53.8 (16.7) | 68.9 (18.8) | 60.8 (18.8) |
| Median (Min, Max) | 56.5 (30.0, 80.0) | 74.0 (39.0, 95.0) | 61.0 (39.0, 95.0) |
| **Brain Weight (g)** | | | |
| Mean (SD) | 1310 (75.5) | 1360 (121) | 1330 (98.3) |
| Median (Min, Max) | 14310 (1210. 1470) | 1320 (1160, 1500) | 1320 (1160, 1500) |
| **APOE Genotype** | | | |
| 2/3 | 1 (12.5%) | 0 (0%) | 1 (6.7%) |
| 3/3 | 5 (62.5%) | 6 (85.7%) | 11 (73.3%) |
| 3/4 | 2 (25.0%) | 1 (14.3%) | 3 (20.0%) |

**Supplementary Methods Table 2:** Demographics of healthy cognitive agers split by lifetime cognition phenotype.

##### *Tissue preparation*

Protocols for post-mortem brain processing were detailed previously [4, 5]. Brain tissues were isolated and processed by freezing for biochemistry and synaptoneurosome preparation and embedding for histopathology (IHC) and array tomography (AT) (Supplementary Methods Fig. 2). Brain regions studied included: primary visual cortex (BA17); middle temporal gyrus (BA20/21); anterior cingulate cortex (BA24); dorsolateral prefrontal cortex (BA46) and posterior hippocampus, all involved in cognitive change during ageing and neurodegenerative diseases [6, 7]. For AT, RNA-seq and proteomics, two of these brain regions BA17 and BA20/21 were investigated, (for AT, ML n=10; HA n=16; AD n=13; and omics ML n=10; HA n=16; AD n=13). BA20/21 is a cortical area involved in working memory and exhibits a larger pathological burden in AD, whereas BA17 is involved in visuo-spatial information processing, and displays a lower pathological load until later disease stages, so is relatively spared during AD [8]. For IHC, all five brain regions were investigated (ML n=6; HA n=13; AD n=6).

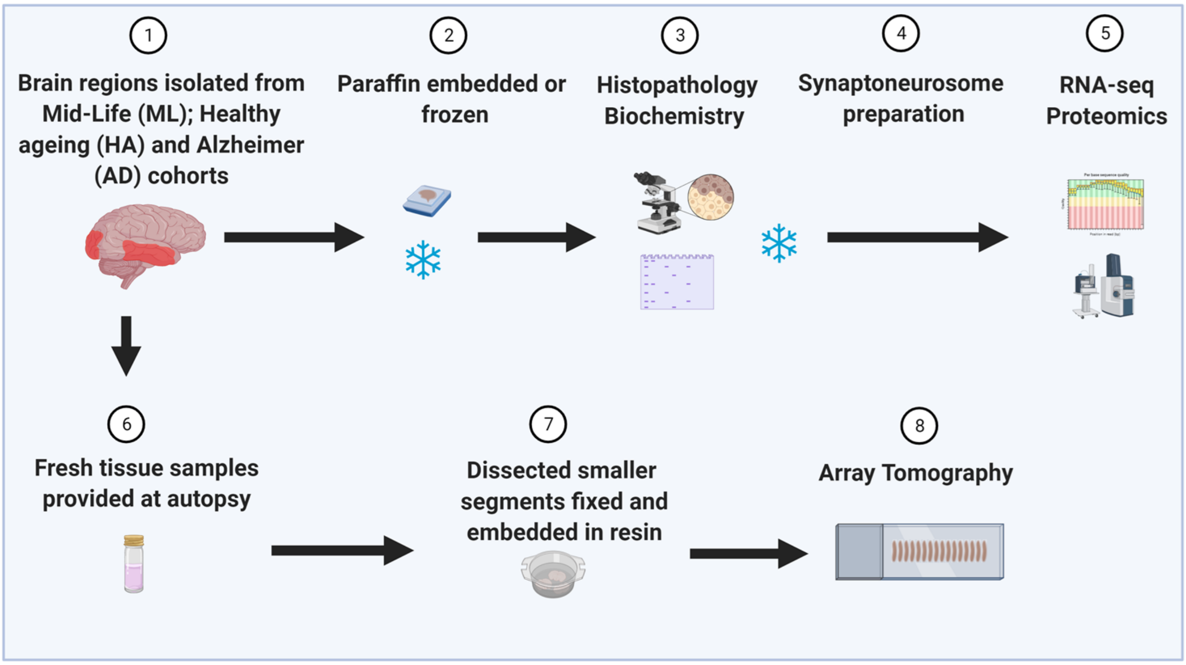


**Supplemental Methods Fig. 2: Summary of post-mortem tissue processing.** Numerous processing techniques employed here allow for in depth insights to brain integrity. Here we show a multi-system approach including histopathological, biochemical, omics and array tomography methods employed in this study. Tissue sampling was predominantly from the left-brain hemisphere. Figure created with BioRender.com.

***Synaptoneurosome preparation***

Both total tissue homogenate and synaptoneurosome fractions were prepared using frozen tissue samples with minor adjustments as previously described [5, 9, 10]. 500mg of was homogenised in a glass-teflon homogeniser with 1ml ice-cold buffer A (25mmol/L HEPES [pH 7.5], 120mmol/L NaCl, 5mmol/L KCl, 1mmol/L MgCl2, and 2mmol/L CaCl2) supplemented with, protease inhibitors (Roche complete mini tablets), phosphatase inhibitors (Merck Millipore) and 1U/µL RNase (Recombinant RNasin Ribonuclease inhibitor 40U/µL). Homogenate was passed through an 80µm nylon net filter (Millipore) to remove tissue debris. A 300µL aliquot was retained and split into two (1:2 ratio) and stored at -80ºC for protein and RNA extraction, respectively. Both fractions are referenced here as total homogenate protein (THp) and/or total homogenate RNA (THr). To prepare filtered synaptoneurosomes the remaining homogenate was passed through a Millex-SV 5µm membrane filter (Millipore) to remove large organelles and nuclei. This aliquot was equally split into two and centrifuged at 1000 × g for 5 minutes. The pellets were washed once with buffer A and centrifuged again yielding two synaptoneurosome pellets. One pellet was stored at -80ºC for synaptoneurosome protein extraction (SNp) and one for synaptoneurosome RNA extraction (SNr). Both THp and SNp aliquots were reconstituted and homogenised with 450µL ice-cold buffer A (100mM Tris-HCL, 4% (w/v) SDS supplemented with protease inhibitor cocktail; Thermo Scientific). Homogenates were centrifuged at 17,000g for 20 minutes. The supernatant was removed and placed in fresh Lo-Bind tubes (Sigma Aldrich) and stored at -80ºC. Protein quantification was carried out using the micro Bicinchoninic acid assay (Pierce, UK) according to manufacturer's guidelines. BCA readings for each brain region and fraction preparation (total homogenate/ synaptoneurosome) were recorded accordingly (DOI File King_et_al_All_data).

***Label-Free quantitative (LFQ) mass spectrometry (MS)***

Sample preparation for LFQ-MS was carried out based on previous proteomic workflows [11, 12]. Protein extracted samples (~60µg; 4% SDS + 100mM Tris-HCL) were processed using a standard S-Trap protocol [13]. Sample digestion was carried out using trypsin enzyme (1:10 to substrate). After digestion, samples were quantified using a fluorometric peptide quantitation assay (Pierce Quantitative Fluorometric Peptide Assay, Thermo Scientific). Samples were reconstituted with 5% Formic Acid (FA) / 10% Acetonitrile (ACN), vortexed, diluted in d$H_2O$ (1:5) then centrifuged (14,500 rpm) for 5 minutes. Approximately 20% of samples were QC tested to ensure samples were free of contaminants (polymers) prior to MS. For QC analysis, 2µl of sample and 8µl 1% FA were mixed, then 5µl was injected onto a Q-Exactive Plus MS system (Thermo Scientific) to assess the level of contamination. QC runs were manually interrogated to determine extent of potential contamination with a pass or fail result. nLC-MS analysis of peptides was performed using a Q-Exactive-HF (Thermo Scientific) mass spectrometer coupled with a dionex Ultimate 3000 RSLCnano (Thermo Scientific). nLC buffers used were as follows: buffer A (0.1% FA in Milli-Q water (v/v)) and buffer B (80% ACN and 0.1% FA in Milli-Q water (v/v)). Aliquots of 1µg of each sample were loaded at 10 µL/ minute onto a trap column (100µm × 2cm, PepMap EasySpray C18 column, 5 µm, 100 Å, Thermo Scientific) equilibrated in buffer A. The trap column was washed for 5 minutes at the same flow rate with buffer A, then was switched in-line with a resolving C18 column (75µm × 50cm, PepMap RSLC C18 column, 2µm, 100 Å, Thermo Scientific). The peptides were eluted from the column at a constant flow rate of 300 nl/ minute with a linear gradient from 2% buffer B to 5% buffer B in 5 minutes, then from 5% buffer B to 35% buffer B in 115 minutes, and then to 98% buffer B within 2 minutes. The column was then washed with 98% buffer B for 15 minutes and re-equilibrated in 2% buffer B for 21 minutes. The column was kept all the time at a constant temperature of 50$^oC$. Q-Exactive HF-X was operated in data dependent positive ionisation mode. The source voltage was set to 3.1 Kv and the capillary temperature was 250$^oC$. A scan cycle comprised MS1 scan (m/z range from 335-1800, with a maximum ion injection time of 50 milliseconds, a resolution of 60,000 and automatic gain control (AGC) value of 3x106) followed by 40 sequential dependant MS2 scans (resolution 7,500 of the most intense ions fulfilling predefined selection criteria (AGC 1x105, maximum ion injection time 50 milliseconds, isolation window of 1.4 m/z, fixed first mass of 120 m/z, NCE/Stepped nce27, spectrum data type: centroid, minimum AGC 2.50X103, exclusion of unassigned, >6 charged precursors, peptide match preferred, exclude isotopes on, dynamic exclusion time of 45s). To ensure mass accuracy, the mass spectrometer was calibrated on the first day that the runs are performed. The raw mass spectrometric data files obtained for each experiment were collated into a single quantitated data set using MaxQuant (version 1.6.0.16) [14] using the Andromeda search engine software [15]. Enzyme specificity was set to trypsin. Parameters used were: (i) variable modifications, methionine oxidation, protein N-acetylation, Phospho (STY), deamidation (NQ); (ii) fixed modifications, cysteine carbamidomethylation; (iii) database: SWISS-Prot-human (20210325); (iv) LFQ: minimum ratio count, 2 (v) MS/MS tolerance: FT-MS 10ppm , FT-MSMS 0.06 Da; (vi) maximum peptide length, 6; (vii) maximum missed cleavages, 2; (viii) maximum of labelled amino acids, 3; and (ix) false discovery rate, 1%. Match Between Runs (MBR) was set to true. LFQ intensities were reported individually for each sample and are given as a relative protein quantitation across all samples. LFQ intensities are represented by a normalised intensity profile generated using algorithms described previously by Cox [16]. Intensities form a matrix with number of samples and number of protein groups as dimensions. Protein differential expression analysis was performed using DEP (Differential Enrichment analysis of Proteomics data; R package version 1.6.1) [17]. In any particular comparison between conditions, a missing value rate threshold was calculated as the maximum of a third of the number of samples in any condition, and any protein with a greater number of missing values than the threshold, in all conditions, was removed. Remaining missing values in the intensity matrix were then imputed using the k-nearest neighbour approach (DOI File King_et_al_All_Omic_data).

***SDS-PAGE and immunoblotting***

SDS-PAGE and immunoblotting was performed as described previously [5, 18]. 20µg of protein from crude total homogenates and synaptoneurosome fractions along with molecular weight marker (Li-Cor) was loaded onto NuPAGE 4-12% Bis-Tris precast polyacrylamide 15 well gel (Invitrogen). Proteins were transferred to iBlot transfer polyvinylidene fluoride (PDVF, Thermofisher) membranes and blocked using Intercept Blocking buffer (Li-Cor). Primary antibodies used for immunoblotting are shown in Supplementary Methods Table 3 and were incubated overnight in blocking buffer. Proteins were detected on an Odyssey system using 680 and 800 IR dye secondary antibodies diluted 1:10,000 in blocking buffer (Supplementary Methods Fig. 3). Total protein stains were performed with Ponceau S and/or REVERT total protein stain as per manufacturer's instructions (Expedeon/Li-Cor).

| Antibody (Ab) | Company | Code/ RRID | Concentration Dilution | Pre-treatment/Secondary Ab |
|---|---|---|---|---|
| **Western Blot** | | | | |
| Synaptophysin | Abcam | Ab8049/ AB_2198854 | 1mg/ml, 1:1000 | Donkey Anti-Mouse; 680RD |
| PSD-95 | Cell Signalling | D27E11/ AB_2292883 | 1mg/ml, 1:1000 | Donkey Anti-Rabbit; 800CW |
| Histone | Abcam | Ab1791/ AB_302613 | 0.1mg/ml, 1:1000 | Donkey Anti-Rabbit; 800CW |
| Clusterin | Abcam | Ab69644/ AB_1267705 | 0.1mg/ml, 1:500 | Donkey Anti-Rabbit; 800CW |
| Stomatin | Abcam | Ab166623 | 0.669mg/ml, 1:500 | Donkey Anti-Rabbit; 800CW |
| Vimentin | Santa Cruz | Sc6260/ AB_628437 | 0.2mg/ml,1:500 | Donkey Anti-Mouse; 680RD |
| **Histopathology** | | | | |
| Amyloid Beta (BA4) | Dako | M087201/ AB_2056966 | 1mg/ml, 1:100 | 98% formic acid |
| CD68 | Dako | M0876/ AB_2074844 | 1ml, 1:100 | Pressure cooker/citric acid |
| GFAP | Dako | Z0334/ AB_10013382 | 1ml, 1:800 | N/A |
| **Array Tomography** | | | | |
| Synaptophysin | Abcam | Ab8049/ AB_2198854 | 1mg/ml, 1:50 | Donkey Anti-Mouse; CY3 |
| PSD-95 | Synaptic systems | 124014/ AB_2619800 | 0.1mg/ml, 1:50 | Donkey Anti-Guinea pig; 488 |
| OC | Merck | Ab2286/ AB_1977024 | 0.1ml, 1:200 | Donkey Anti-Rabbit; 405 |
| Total Tau | R&D Systems | AF3494/ AB_573209 | 0.1mg/ml, 1:50 | Donkey Anti-Goat; 647 |
| **Immunocytochemistry** | | | | |
| OCT3/4 (POU5F1) | Abcam | Ab181557/AB_2687916 | 3µg/ml, 1:250 | Donkey Anti-Rabbit; 488 |
| SSEA4 | Abcam | Ab16287/AB_778073 | 15.5µg/ml, 1:300 | Donkey Anti-Mouse; 568 |
| SOX2 | R&D Systems | AF2018/AB_355110 | 0.2µg/ml, 1:1000 | Donkey Anti-Goat; 647 |
| OTX1/2 | Abcam | Ab181557/AB_776930 | 1µg/ml, 1:1000 | Donkey Anti-Rabbit; 488 |
| Nestin | Abcam | Ab6320/AB_308832 | 5µg/ml, 1:100 | Donkey Anti-Mouse; 568 |
| MAP2 | Synaptic systems | 188004/AB_2138181 | 1:1,000 | Goat Anti-Guinea Pig; 647 |
| CUX1 | Atlas Antibodies | HPA003317/AB_2666891 | 1µg/ml, 1:1000 | Donkey Anti-Rabbit; 488 |
| GFAP | Abcam | Ab10062/AB_296804 | 4µg/ml, 1:500 | Donkey Anti-Mouse; 568 |
| SATB2 | Abcam | Ab92446/AB_10563678 | 14.4µg/ml, 1:100 | Donkey Anti-Rabbit; 488 |
| Homer1 | Abcam | Ab211415/AB_2094490 | 1.5µg/ml, 1:500 | Donkey Anti-Rabbit; 488 |
| 6E10 | BioLegend | 803001/AB_2564653 | 10µg/ml, 1:100 | Donkey Anti-Mouse; 568 |
| DAPI | Sigma-Aldrich | D9542/AB_181557 | 10mg, 1:10000 | N/A |

**Supplementary Methods Table 3:** Primary and secondary antibody details for western blotting, histopathology, array tomography and iPSC model.

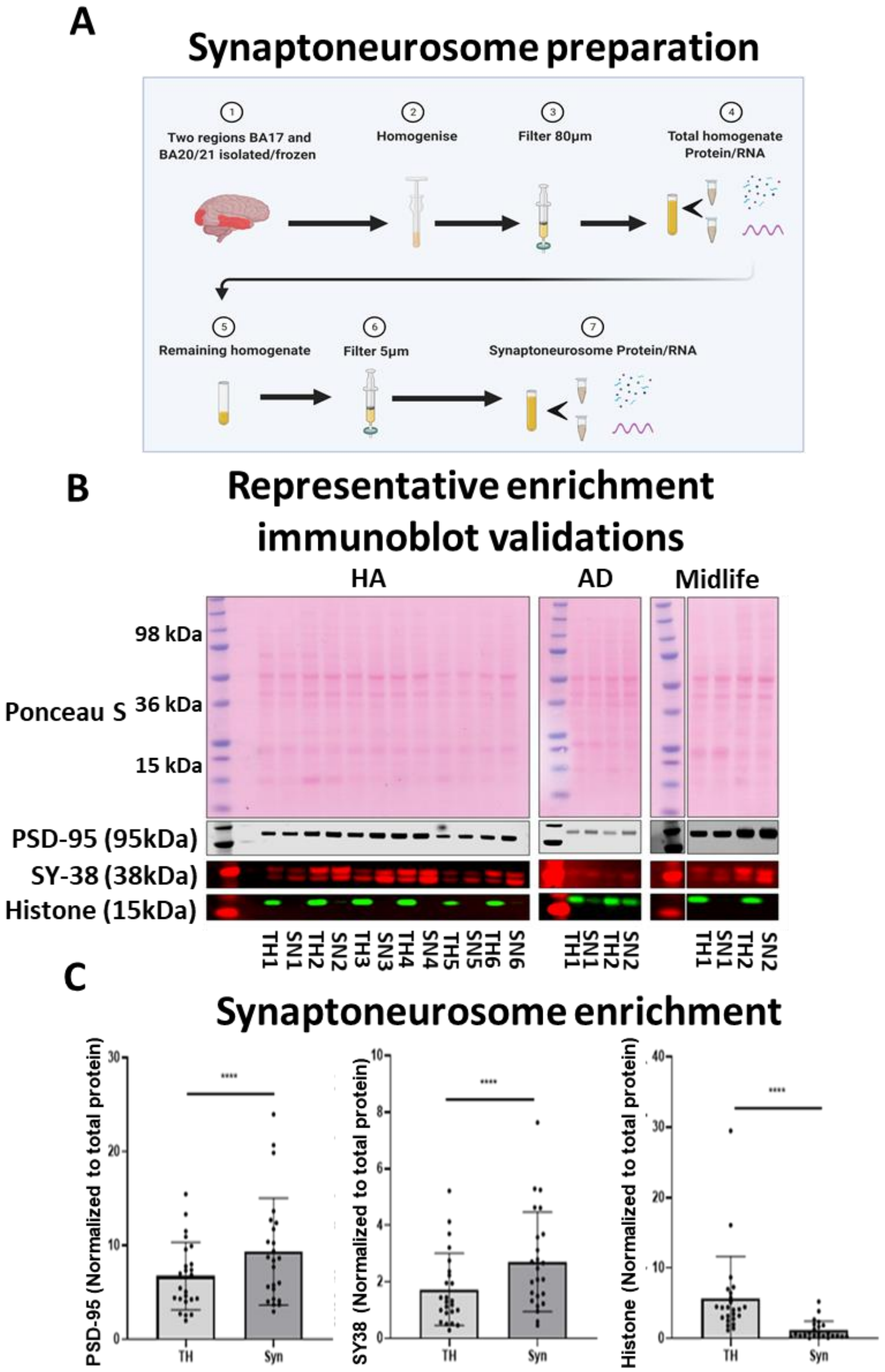
A
Synaptoneurosome preparation
1
Two regions BA17 and BA20/21 isolated/frozen
2
Homogenise
3
Filter 80µm
4
Total homogenate Protein/RNA
5
Remaining homogenate
6
Filter 5µm
7
Synaptoneurosome Protein/RNA
B
Representative enrichment immunoblot validations
HA
AD
Midlife
98 kDa
Ponceau S
36 kDa
15 kDa
PSD-95 (95kDa)
SY-38 (38kDa)
Histone (15kDa)
TH1 SN1 TH2 SN2 TH3 SN3 TH4 SN4 TH5 SN5 TH6 SN6
TH1 SN1 TH2 SN2
TH1 SN1 TH2 SN2
C
Synaptoneurosome enrichment
PSD-95 (Normalized to total protein)
SY38 (Normalized to total protein)
Histone (Normalized to total protein)
****
TH
Syn

**Supplementary Methods Fig. 3:** Synaptoneurosome preparation and quality control. **A** Step-by-step flowchart summarizing the steps involved in synaptoneurosome preparation from two brain regions namely primary visual cortex and middle temporal gyrus from HA, AD and mid-life brains. **B** Representative images from western blotting. TH refers to total homogenate and SN to synaptoneurosome preparations. The imaged western blots were cropped to only show the diagnostic bands that were used for densitometry analysis. **C** Analysis of immunolabelled blots from HA cases confirm synaptic enrichment and histone exclusion. Paired t tests; PSD-95 p value $<0.0001$ ($t=7.106$, $df=23$); SY38 p value $<0.0001$ ($t=9.380$, $df=23$); Histone p value $<0.0001$ ($t=13.40$, $df=23$), $n=24$, n=individual samples. Data presented in graph form as Arbitrary Units (AU).

##### *RNA extraction and analysis*

mRNA was extracted from both THr and SNr samples isolated from synaptoneurosome preparations using the RNeasy Plus Micro Kit (Qiagen) according to manufacturer's instructions. RNA was assessed for quality (Agilent, TapeStation) and quantity (Invitrogen, Qubit) before library preparation (DOI File King_et_al_All_data). Illumina libraries were prepared from 1µg of RNA using TruSeq mRNA Sample Prep Kit as per the manufacturer's recommendations. Illumina sequencing was carried out on a NovaSeq platform using 50 base paired-end reads. Raw reads were processed using RTA 1.17.21.3 and bcl2fastq v2.20 (Illumina). Reads were mapped to human (hg38) reference genomes using version 2.7.0 of the STAR RNA-seq aligner [19]. A table of per-gene read counts was generated from the mapped reads with featureCounts version 1.6.3 [20], using gene annotations from Ensembl version 96. Differential expression analysis was then performed using DESeq2 (R package version 1.24.0) [21] followed by gene set analysis using Camera [22] from the limma R package (version 3.40.6) [23] and Gene Ontology enrichment analysis using topGO (R package version 2.36; (https://rdrr.io/bioc/topGO/); (DOI File King_et_al_All_Omic_data). To validate a subset of sequencing results, qPCR was performed in an CFX96 Real-time system (Bio-Rad) using BRYT Green Dye (Promega) and the GoTaq 1-Step RT-qPCR kit (Promega) according to manufacturer's instructions. Briefly, RNA (equivalent to 100ng) was mixed with concentrated GoTaq qPCR Master Mix and GoScript RT mix, appropriate forward and reverse primers (250nM) and made to a final volume of 20µl with RNase-free water. Technical replicates as well as no template controls (NTC's) were included in each run. For post-mortem tissue experiments, gene expression levels were normalized in all cases to *KCP, CCR9 and EN2* housekeeping controls (selected based on coefficient of variation values from RNA-seq dataset). For iPSC-neuron experiments, *GAPDH* and *RPLP1* were the housekeeping controls used. Primer sequences used for validation purposes are shown in Supplemental Methods Table 4.

| Primer | Sequence |
|---|---|
| KCP | Forward Sequence - ACCTGTCTTGGAGGCTTCGTGA |
| | Reverse Sequence - AGGCAGTAGTGACGCCATGGTA |
| CCR9 | Forward Sequence - TCGTGGTCATGGCTTGCTGCTA |
| | Reverse Sequence - AAGACGGTCAGGACAGTGATGG |
| EN2 | Forward Sequence - CGCGCAGCCCATGCTCTGGC |
| | Reverse Sequence - GCTTGTCCTCTTTGTTCGGGTTC |
| RRAS | Forward Sequence - CTGCTGGTGTTCGCCATTAACG |
| | Reverse Sequence - GATCTGCCTTGTTCCCGACCAA |
| ITPR1 | Forward Sequence - GTGACAGGAAACATGCAGACTCG |
| | Reverse Sequence - CAGCAGTTGCACAAAGACAGGC |
| GAD2 | Forward Sequence - TGTCCAGGAAGCACCGCCATAA |
| | Reverse Sequence - TCCTTGACGAGAATGGCAGAGC |
| SNAP-25 | Forward Sequence - CGTCGTATGCTGCAACTGGTTG |
| | Reverse Sequence - GGTTCATGCCTTCTTCGACACG |
| TM4SF1 | Forward Sequence - GGCTACTGTGTCATTGTGGCAG |
| | Reverse Sequence - ACTCGGACCATGTGGAGGTATC |
| SYT1 | Forward Sequence - GCTGACTGTTGTCATTCTGGAGG |
| | Reverse Sequence - CTTCAGCCTCTTACCATTCTG |
| GAPDH | Forward Sequence - CCAAGGTCATCCATGACAAC |
| | Reverse Sequence - ACAGTCTTCTGGGTGGCAGT |
| RPLP1 | Forward Sequence - AGCCGGTGTAAATGTTGAGC |
| | Reverse Sequence - CAGATGAGGCTCCCAATGTT |

**Supplementary methods Table 4:** Sequences of qPCR primer pairs.

***Histopathology***

Brain tissue processing for neuropathology has been described previously [5, 24]. Fresh post-mortem tissue blocks were fixed in 10% formalin, dehydrated in an ascending series of alcohol (70-100%) then finally paraffin embedded. Tissue sections were cut on a Leica microtome at 4μm thickness and processed for immunohistochemistry using the Novolink Polymer Detection Kit (Leica). The chromogen used for visualization was 3,3'-diaminobenzidine (DAB) with 0.05% hydrogen peroxide as substrate. Antibody information is shown in Supplementary Methods Table 3. Tissues were counterstained with haematoxylin for 30 seconds to visualise cell nuclei. Representative histopathological images are shown in Supplementary Fig. 1. All immunolabelled sections were assessed blind to case information and stain burdens calculated using Stereo Investigator (MBF Bioscience 2019). Cortical grey matter was outlined in each section at 1.5X objective, then immuno-positive objects were identified at X5 objective using an automated colour-based thresholding algorithm. The area of immuno-positive cortex was expressed as a percentage area of total cortex in each brain region, calculated using the Neurolucida Explorer (MBF Bioscience 2019) software.

***Array Tomography***

Fresh brain samples were fixed in 4% paraformaldehyde, dehydrated, and embedded in LR white resin as previously described [5, 25, 26]. Ribbons of ultrathin serial sections were cut on an ultramicrotome (Leica) with a histo jumbo diamond knife (Diatome). AT ribbons were stained with immunofluorescence and imaged using an AxioImager Z2 with an 63x 1.4 NA objective (Supplementary Fig. 1). Images were acquired in Zen software 2.1 (Zeiss) and included a series of serial sections from the same fixed location. If Aβ plaques were present, serial images were taken of the plaque and of a plaque-free area in the same cortical region. Individual serial images were combined into stacks using Fiji (Image J). Images were aligned (via affine and rigid registration, using puncta from the synaptophysin channel as the reference), segmented (done by thresholding stacks using a semi-automatic local threshold based on mean intensity values) and objects detected in three dimensions using custom Matlab script. The co-localization between channels was calculated using custom Python script, to determine the percentage of paired synaptic terminals and the percentage of synapses co-localized with either Aβ or total tau. All custom software can be downloaded from GitHub at: https://github.com/arraytomographyusers/Array_tomography_analysis_tool and https://github.com/lewiswilkins/Array-Tomography-Tool. 3D reconstructions of representative images were created with IMARIS software version 9.8.

***iPSC culture***

Peripheral blood mononuclear cells (PBMCs) from the LBC1936 cohort were reprogrammed to iPSCs using non-integrating oriP/EBNA1 backbone plasmids expressing six iPSC reprogramming factors and lines were grouped into LCR (n=4) and LCD (n=2) categories, as described previously [27]. The iPSCs were maintained on 1:100 geltrex (Thermo, A1413302), fed daily with Essential 8 media (Thermo, A1517001 and A15171-01), and passaged with 0.1% EDTA (Thermo, 15575-020). The iPSCs were differentiated to glutamatergic cortical neurons by dual SMAD inhibition, following a protocol adapted from Shi et al. [28]. For neural induction, confluent iPSCs were fed daily with induction media N2B27 media, with 10μM SB431542 (Tocris, 1614) and 1μM dorsomorphin (R&D Systems, 3093/10) for 12 days. Neuroepithelium was passaged mechanically onto plates coated with 1:200 Geltrex and 10μg/mL laminin (Merck L2020-1MG) and fed with N2B27 every two days. Neural precursor cells (NPCs) were passaged with accutase (Thermo, A11105-01) at days 18, 25 and 30-post induction. At final passage, NPCs were seeded at a density of 50,000 cells per cm3 into 24 well plates containing glass coverslips coated with 20μg/mL poly-L-ornithine (Merck P4957), 10μg/mL laminin, 10μg/mL fibronectin (Merck, F2006), and 250μg/mL matrigel (Corning 354230). Between days 7-21 post-final passage, maturing neurons were fed N2B27 with 10μM forskolin (Tocris, 1099) every three-four days. From day 21 onwards, maturing neurons were fed with N2B27 with 5ng/ml BDNF (R&D Systems, 248-BD) and GDNF (R&D Systems, 212-GD) every three-four days. Characterisation was conducted 60 days post-final passage. Generation and immunodepletion of human brain homogenate for treating iPSC derived neurons was conducted following a protocol adapted from Hong et al. [29]. Human superior temporal gyrus and temporal pole brain regions were homogenised manually with a razorblade, and collected in a low protein binding 15mL tube (Thermo, 30122216) containing 10mL 1X artificial CSF (pH 7.4) supplemented with 1x cOmplete mini EDTA-free protease inhibitor cocktail tablet (Merck, 11836170001) per 10mL, per 2g of tissue. The solution was mixed for 30 minutes to extract soluble proteins, then centrifuged at 2000 RCF for 10 minutes to remove large, insoluble debris. The supernatant was transferred to ultracentrifuge tubes (Beckman, 355647) and then centrifuged at 200,000 RCF for 110 minutes. The resulting supernatant, a homogenate fraction containing soluble Aβ forms, was then transferred to a Slide-A-Lyser G2, 2K MWCO 15mL dialysis cassette (Thermo, 87719) and dialysed in 1X aCSF with magnetic stirring for three days to remove salts from the homogenate. The 1X aCSF was exchanged every 24 hours. Dialysed brain homogenate was divided into two equal portions in low protein binding 15mL tubes. Protein A Agarose (PrA) beads (Thermo, 20334) were washed three times in 1X aCSF. 30µL of washed beads were added per 1mL of homogenate. To create Aβ- treatment samples, Aβ was immunodepleted from one homogenate fraction by adding 20µL 4G8 antibody (Biolegend, 800711) per 1mL of homogenate. To create Aβ+ treatment samples, the other homogenate fraction was 'mock-immunodepleted' by adding 20µL of mouse serum per 1ml of homogenate. Homogenate was then incubated for 24 hours with gentle mixing. Next, homogenate was centrifuged at 2500 RCF for 5 minutes to remove the beads, and the supernatant collected. The process of adding beads and antibody/serum to homogenate was repeated twice more. After the third centrifugation step, PrA beads alone were added to both Aβ+ and Aβ- homogenate, incubated for two hours with gentle mixing, and then centrifuged at 2500 RCF for 5 minutess to clear any remaining antibody. Homogenate from each portion was aliquoted at 0.5mL into 1.5mL low protein binding Eppendorf's (Thermo, 0030108116) and stored at -80°C. Concentration of Aβ1-42 in Aβ+ and Aβ- homogenate was quantified by sandwich ELISA (WAKO, 296-64401), according to manufacturer instructions. Cells cultured on glass coverslips were washed with 1X D-PBS (Thermo), fixed with 4% formalin (Polysciences, 04018-1) for 15 minutes, and washed in 1X D-PBS three times. For staining, cells were permeabilised and blocked for 1 hour with D-PBS containing 0.3% Triton-X (Merck, X100-500ML) and 10% goat serum (Merck, 566380-10ML). Cells were incubated overnight at 4°C with primary antibody for appropriate markers, washed in D-PBS, then incubated with secondary antibodies for one hour (Supplementary Methods Table 3). Cells were then washed once with D-PBS, incubated for 10 minutes with DAPI diluted in D-PBS containing 0.3% Triton-X, then washed in D-PBS. Coverslips were mounted on glass microscope slides (VWR, 631-0847) with mounting media (Merck, 345789) and stored at 4°C until use. Coverslips were imaged on a Leica TCS confocal microscope with an oil immersion 63x objective. Five fields per coverslip containing

MAP2 staining were randomly selected for imaging and image stacks acquired through the thickness of the cell layer (0.3µm per step). Image stacks were processed using custom software to segment staining, calculate the density of homer1 co-localized along MAP2 positive processes. All image analysis scripts are freely available at https://github.com/Spires-Jones-Lab. 3D reconstructions of representative images were created with IMARIS software version 9.8.

***Statistics***

Group comparisons including variables Cohort (or cognitive status), Sex, PMI, APOE status, Plaque present/absent) were analysed using Linear mixed effects models including case as a random effect to account for multiple measures per case. Specifically for array tomography, tissue sample was nested within case as a random effect and experimenter was included as a random effect as more than one person collected the data. To assess whether data met assumptions of normal distribution, model residuals were visually inspected via QQ plot, coupled with Shapiro-Wilk testing. Where data failed to meet model assumptions (Shapiro-Wilk $p < 0.05$), data were transformed via Tukey's Ladder of Power. Type III ANOVA with Satterthwaite correction were performed on the linear mixed effects models to assess the significance of main effects. For subsequent pairwise comparisons between experimental groups, this was followed by Tukey-corrected post-hoc testing. All analyses were performed using R Studio [30] (R 4.4.1) and the scripts and full statistical results can be found in Supplementary data with associated spreadsheets available on the Edinburgh DataShare repository.

1. *The trend of Scottish intelligence: a comparison of the 1947 and 1932 surveys of the intelligence of eleven-year-old pupils*. The trend of Scottish intelligence: a comparison of the 1947 and 1932 surveys of the intelligence of eleven-year-old pupils. 1949, Oxford, England: University London Press. xxviii, 151-xxviii, 151.
2. Deary, I.J., et al., *The Lothian Birth Cohort 1936: a study to examine influences on cognitive ageing from age 11 to age 70 and beyond.* BMC Geriatr, 2007. **7**: p. 28.
3. Deary, I.J., Whalley, L. J., & Starr, J. M. , *A Lifetime of Intelligence: Follow-Up Studies of the Scottish Mental Surveys of 1932 and 1947*. 2009: Washington, DC: American Psychological Association. 285.
4. Samarasekera, N., et al., *Brain banking for neurological disorders.* Lancet Neurol, 2013. **12**(11): p. 1096-105.
5. Henstridge, C.M., et al., *Post-mortem brain analyses of the Lothian Birth Cohort 1936: extending lifetime cognitive and brain phenotyping to the level of the synapse.* Acta Neuropathol Commun, 2015. **3**(1): p. 53.
6. Keller, J.N., *Age-related neuropathology, cognitive decline, and Alzheimer's disease.* Ageing Research Reviews, 2006. **5**(1): p. 1-13.
7. Cui, J.G., et al., *Expression of inflammatory genes in the primary visual cortex of late-stage Alzheimer's disease.* Neuroreport, 2007. **18**(2): p. 115-9.
8. Ltd, T.R., *Cortical Functions Reference Manual.* Trans Cranial Technologies, 2012.
9. Tai, H.C., et al., *The synaptic accumulation of hyperphosphorylated tau oligomers in Alzheimer disease is associated with dysfunction of the ubiquitin-proteasome system.* Am J Pathol, 2012. **181**(4): p. 1426-35.
10. Hollingsworth, E.B., et al., *Biochemical characterization of a filtered synaptoneurosome preparation from guinea pig cerebral cortex: cyclic adenosine 3':5'-monophosphate-generating systems, receptors, and enzymes.* J Neurosci, 1985. **5**(8): p. 2240-53.
11. Sticker, A., et al., *Robust Summarization and Inference in Proteome-wide Label-free Quantification.* Molecular & Cellular Proteomics, 2020. **19**(7): p. 1209-1219.
12. Tebbe, A., et al., *Systematic evaluation of label-free and super-SILAC quantification for proteome expression analysis.* Rapid Communications in Mass Spectrometry, 2015. **29**(9): p. 795-801.
13. HaileMariam, M., et al., *S-Trap, an Ultrafast Sample-Preparation Approach for Shotgun Proteomics.* J Proteome Res, 2018. **17**(9): p. 2917-2924.
14. Cox, J. and M. Mann, *MaxQuant enables high peptide identification rates, individualized p.p.b.-range mass accuracies and proteome-wide protein quantification.* Nature Biotechnology, 2008. **26**(12): p. 1367-1372.
15. Cox, J., et al., *Andromeda: a peptide search engine integrated into the MaxQuant environment.* J Proteome Res, 2011. **10**(4): p. 1794-805.
16. Cox, J., et al., *Accurate proteome-wide label-free quantification by delayed normalization and maximal peptide ratio extraction, termed MaxLFQ.* Mol Cell Proteomics, 2014. **13**(9): p. 2513-26.
17. Zhang, X., et al., *Proteome-wide identification of ubiquitin interactions using UbIA-MS.* Nat Protoc, 2018. **13**(3): p. 530-550.
18. Hesse, R., et al., *Comparative profiling of the synaptic proteome from Alzheimer's disease patients with focus on the APOE genotype.* Acta Neuropathol Commun, 2019. **7**(1): p. 214.
19. Dobin, A., et al., *STAR: ultrafast universal RNA-seq aligner.* Bioinformatics, 2013. **29**(1): p. 15-21.
20. Liao, Y., G.K. Smyth, and W. Shi, *featureCounts: an efficient general purpose program for assigning sequence reads to genomic features.* Bioinformatics, 2014. **30**(7): p. 923-30.
21. Love, M.I., W. Huber, and S. Anders, *Moderated estimation of fold change and dispersion for RNA-seq data with DESeq2.* Genome Biology, 2014. **15**(12): p. 550.
22. Wu, D. and G.K. Smyth, *Camera: a competitive gene set test accounting for inter-gene correlation.* Nucleic Acids Research, 2012. **40**(17): p. e133-e133.
23. Ritchie, M.E., et al., *limma powers differential expression analyses for RNA-sequencing and microarray studies.* Nucleic Acids Res, 2015. **43**(7): p. e47.

24. Tzioras, M., et al., *Assessing amyloid-β, tau, and glial features in Lothian Birth Cohort 1936 participants post-mortem.* Matters, 2017.
25. Kay, K.R., et al., *Studying synapses in human brain with array tomography and electron microscopy.* Nat Protoc, 2013. **8**(7): p. 1366-80.
26. Kurucu, H., et al., *Inhibitory synapse loss and accumulation of amyloid beta in inhibitory presynaptic terminals in Alzheimer's disease.* Eur J Neurol, 2021.
27. Toombs, J., et al., *Generation of twenty four induced pluripotent stem cell lines from twenty four members of the Lothian Birth Cohort 1936.* Stem cell research, 2020. **46**: p. 101851-101851.
28. Shi, Y., P. Kirwan, and F.J. Livesey, *Directed differentiation of human pluripotent stem cells to cerebral cortex neurons and neural networks.* Nature Protocols, 2012. **7**(10): p. 1836-1846.
29. Hong, W., et al., *Diffusible, highly bioactive oligomers represent a critical minority of soluble Aβ in Alzheimer's disease brain.* Acta neuropathologica, 2018. **136**(1): p. 19-40.
30. R Core Team, *R: a language and environment for statistical computing*. 2017.
